# Early sodium channel blocker use improves seizures and neurodevelopment in *KCNQ2*-related disorders

**DOI:** 10.64898/2026.02.10.26345394

**Authors:** Charissa Millevert, Megan Hairabedian, Johannes Lemke, Steffen Syrbe, Eugenia Roza, Raluca Ioana Teleanu, Laura Licchetta, Duccio M Cordelli, Francesca Bisulli, Trine B Hammer, Magdalena Krygier, Marta Pietruszka, Maria Mazurkiewicz-Bełdzińska, Safa Mete Dagdas, Pinar Gencpinar, Carmen Fons, Dídac Casas-Alba, Edward C Cooper, Maurizio Taglialatela, Béatrice Desnous, Nathalie Villeneuve, Anne Lépine, Stéphane Auvin, Cyril Mignot, Dorothée Ville, Anne de Saint Martin, Claire Bar, Caroline Hachon le Camus, Laurent Villard, Laurence Chaton, Patrick Van Bogaert, Jérémie Lefranc, Gaëtan Lesca, Silvia Napuri, Mathieu Kuchenbuch, Caroline Perriard, Blandine Dozieres, Bénédicte Héron, Annie Ting-Gee Chiu, Ingrid E Scheffer, KCNQ2 study group, Mathieu Milh, Sarah Weckhuysen

**Affiliations:** VIB Center for Molecular Neurology, University of Antwerp, 2610, Antwerp, Belgium; University Hospital of Antwerp, Dept. of Neurology, 2650, Antwerp, Belgium; Faculty of Medicine and Health Science, University of Antwerp, 2610, Antwerp, Belgium; Pediatric Neurology Department, Pediatric Hospitals of Nice, Lenval University Hospital Center, 06200, Nice, France; Institute of Human Genetics, University of Leipzig, 04103 Leipzig, Germany; Department of Pediatrics I, University Hospital Heidelberg, 69120 Heidelberg, Germany; Carol Davila University of Medicine and Pharmacy, Clinical Neurosciences Department, Pediatric Neurology, 050474, Bucharest, Romania; Dr. Victor Gomoiu Children’s Hospital, 022102, Bucharest, Romania; IRCCS Institute of Neurological Sciences of Bologna, Full Member of the European Reference Network for Rare and Complex Epilepsies (EpiCARE), 40139 Bologna, Italy; Department of Medicine and Surgery, University of Bologna, 40126, Bologna, Italy; Department of Biomedical and NeuroMotor Sciences, Alma Mater Studiorum – University of Bologna, 40127, Bologna, Italy; Department of Epilepsy Genetics and Personalized Medicine, Danish Epilepsy Centre, 4293, Dianalund, Denmark; Department of Clinical Genetics, Center of Diagnostics. Copenhagen University Hospital - Rigshospitalet, 2100, Copenhagen, Denmark; Department of Developmental Neurology, Medical University of Gdansk, 80-211 Gdansk, Poland; Department of Pediatric Neurology, Izmir City Hospital, Refik Sevket Ince Mh. 2148/11 Sk. No:1/11, 35530, Bayrakli, Izmir, Turkey; Department of Pediatric Neurology, Izmir Katip Celebi University, Balatcik, Havaalani Sosesi Cd. No:33/2, 35610, Cigli, Izmir, Turkey; Early Onset and genetic Epilepsy Unit, Neurology and Neurophysiology Department, Hospital Sant Joan de Déu, ERN-Epicare Coordinator Centre; Complex Epilepsies Research Group, Institut de Recerca de Sant Joan de Déu. 08950 Esplugues de Llobregat, 08950, Barcelona, Spain; Clinical Genetics Department and Institut de Recerca Sant Joan de Déu, Hospital Sant Joan de Déu, Esplugues de Llobregat, 08950, Spain; Departments of Neurology, Neuroscience, and Molecular and Human Genetics, Baylor College of Medicine, Houston, TX 77030; Section of Pharmacology, Dept. of Neuroscience, University of Naples “Federico II”, 80131 Naples, Italy; Aix-Marseille University, APHM, Department of Pediatric Neurology, Timone Children’s Hospital, 13005, Marseille, France; Autisme Resource Center, APHM, hôpital Sainte Marguerite 13009 Marseille; AP-HP, Pediatric Neurology Department, Reference Center for Rare Epilepsies, Member of ERN Epicare, Robert Debré University Hospital, 75019 Paris, France; Institut Hospitalo-Universitaire Robert-Debré du Cerveau de l’Enfant, 75019 Paris, France; Université Paris-Cité, INSERM Neuro-Diderot, 75019 Paris, France; Institut Universitaire de France (IUF), 75000 Paris, France; Department of Genetics and Reference Center for Rare Intellectual Disability, Pitié-Salpêtrière Hospital, AP-HP, 75013 Paris, France; University Hospitals of Lyon (HCL), Pediatric Neurology Department, and reference Center of rare epilepsies Lyon, 69500, Bron, France; Department of Child Neurology, University Hospital of Strasbourg (CHU Strasbourg), 67000 Strasbourg, France; Department of Pediatric Neurology, Reference Center for Rare Epilepsies, CHU Bordeaux, 33076, Bordeaux, France; CNRS, INCIA, UMR 5287, NRGen Team, Univ.Bordeaux, 33000, Bordeaux, France; CHU Toulouse, Department of Pediatric Neurology, Toulouse Children’s Hospital, 31059, Toulouse, France; Aix Marseille Univ, Inserm, MMG, U1251, 13385, Marseille, France; APHM, Medical Genetics, Marseille University Hospital, 13005, Marseille, France; Clinical Neurophysiology Department, Lille University Hospital, 59037, Lille, France; Department of Pediatric Neurology, University Hospital of Angers (CHU Angers), 49100 Angers, France; Department of Pediatrics, Morvan University Hospital (CHU Morvan), 29200 Brest, France; Department of Medical Genetics, Member of ERN EpiCARE, University Hospitals of Lyon, 69500, Bron, France; Laboratoire Physiopathologie et Génétique du Neurone et du Muscle, Neuromyogene Institute, CNRS UMR 5261 -INSERM U1315, Université de Lyon – Université Claude Bernard Lyon 1, 69100, Villeurbane, France; Department of Pediatrics, Rennes University Hospital, 35033, Rennes, France; Department of Pediatric Neurology, Nancy University Hospital (CHU Nancy), 54000 Nancy, France; Department of Pediatric Neurology, Robert-Debré University Hospital (AP-HP), 75019 Paris, France; Department of Pediatric Neurology, Armand Trousseau Hospital, and FHU I2D2, APHP, Sorbonne University, 75012, Paris, France; Epilepsy Research Centre, Department of Medicine, University of Melbourne, Austin Health, Melbourne, 3084, Victoria, Australia; Bladin-Berkovic Comprehensive Epilepsy Program, Department of Neurology, Austin Health, Melbourne, 3084, Victoria, Australia; Florey Institute and Murdoch Children’s Research Institute, Melbourne, 3052, Victoria, Australia; Department of Paediatrics, University of Melbourne, Royal Children’s Hospital, Melbourne, 3052, Victoria, Australia; INMED, INSERM U1249, Luminy Scientific Park, 13009, Marseille, France

**Keywords:** neonatal epilepsy, channelopathies, genetic epilepsy, developmental and epileptic encephalopathy, precision medicine, antiseizure medication

## Abstract

**Background:** Pathogenic *KCNQ2* variants are the most common genetic cause of neonatal-onset epilepsies, with phenotypes ranging from self-limited (familial) neonatal epilepsy (SeL(F)NE) to severe developmental and epileptic encephalopathy (KCNQ2-DEE). Sodium channel blockers (SCBs) have shown promise for seizure control in these disorders, but their impact on neurodevelopmental outcomes and possible relationship with timing of initiation remain incompletely understood.

**Methods:** We leveraged a large, multicentre international cohort comprising 282 individuals with *KCNQ2* pathogenic variants to retrospectively assess the effectiveness of antiseizure medications (ASMs), particularly SCBs, on seizure control and neurodevelopment. Individuals were grouped according to the predicted variant-specific functional effects: loss-of-function (LOF) variants known to be associated with SeL(F)NE or DEE respectively, and gain-of-function (GOF) variants. Epilepsy course, ASM effectiveness, and neurodevelopmental milestones were systematically collected and analysed.

**Results:** SCBs, especially carbamazepine (CBZ) and oxcarbazepine (OXC), emerged as the most effective ASMs in both LOF groups. In LOF *KCNQ2*-DEE, early SCB initiation within the first month of life was associated with significantly more favourable neurodevelopmental trajectories, including higher rates of achievement of major motor milestones. Early seizure freedom itself was a strong predictor of improved neurodevelopment, with the positive effect of SCBs likely mediated by their ability to control seizures. However, considerable phenotypic variability persisted, with some individuals experiencing severe impairment despite early seizure control and SCB initiation. Variant severity and possible genetic modifiers likely contribute to this heterogeneity, underscoring the need for precision therapies beyond nonspecific ASM approaches.

**Conclusion:** Our results strongly support the use of SCBs as first-line therapy in (LOF) *KCNQ2*-DEE and SeL(F)NE due to their high effectiveness. Moreover, SCBs appear most beneficial when initiated during the neonatal period, with earlier treatment linked to earlier seizure offset and better developmental outcomes. These results highlight the importance of early genetic diagnosis and timely SCB therapy, and support CBZ or OXC as first-line agents. We however emphasise that early treatment is not universally transformative, and further work, including exploration of targeted therapies but also standardised neurodevelopmental assessments, is needed to optimise long-term outcomes in this heterogeneous population.

## Introduction

*KCNQ2* pathogenic variants are the most frequent genetic cause of neonatal-onset genetic epilepsy.^1^ It encodes the Kv7.2 subunit of the voltage-gated Kv7 potassium channel, which contributes to the generation of the M-current, a critical regulator of neuronal excitability in the developing and mature central and peripheral nervous system.^2,3^

Pathogenic variants in *KCNQ2* cause a range of epilepsy phenotypes, with autosomal dominant inheritance and with disease severity correlating with their biophysical impact on the M-current density, as shown by *in vitro* electrophysiology studies.^4,5^ Loss-of-function (LOF) variants, such as large deletions including the whole gene, truncating variants or certain missense variants, reducing Kv7.2 channel function by up to 50% are associated with self-limited (familial) neonatal epilepsy (SeL(F)NE), typically characterised by self-limiting neonatal seizures and normal neurodevelopment (OMIM: 121200).^1^ In contrast, *KCNQ2*-related developmental and epileptic encephalopathy (*KCNQ2*-DEE, OMIM: 613720) is caused by dominant-negative missense or in-frame deletion variants that suppress channel function more severely (>50% reduction in M-current density),^6^ leading to neonatal-onset seizures and moderate to profound developmental impairments.^7–10^ Rare gain-of-function (GOF) variants increase M-current density and lead to neurodevelopmental impairments without neonatal seizures, although epilepsy may emerge later in life (OMIM: 613720).^11,12^ Despite relatively good phenotype-genotype correlations, phenotypic variability with variable degrees of both cognitive impairment and seizure severity is described.^7,13^ This observation points towards the existence of genetic and/or environmental modifiers, such as the use of certain anti-seizure medications (ASMs) as seen in other genetic DEEs.^14–16^

Both the underlying pathogenic variant, the seizures and/or electroencephalogram (EEG) abnormalities likely contribute to early developmental impairment in *KCNQ2*-DEE.^2,17^ Therefore, timely and effective seizure control might be essential not only for acute symptom management but also for optimising developmental outcomes. Sodium channel blockers (SCBs), such as phenytoin (PHT), carbamazepine (CBZ) and oxcarbazepine (OXC), have shown efficacy in controlling *KCNQ2*-related seizures in both SeL(F)NE and DEE in some retrospective studies.^18–20^ In recognition of emerging genotype-treatment correlations, the International League Against Epilepsy (ILAE) Task Force now recommends phenobarbital as the first-line ASM in neonates, except when a genetic channelopathy is suspected, in which case SCBs are preferred.^21^ Despite these recommendations, SCBs remain underutilized in routine neonatal care.^22^

A small case series suggested that early and effective treatment of seizures in *KCNQ2*-DEE may also positively affect neurodevelopmental outcome.^19^ Additionally, a recent study in a knock-in mouse model of *KCNQ2*-DEE showed that early and long-term SCB treatment not only improved seizures but also enhanced cognitive abilities.^23^ However, it remains unclear to what extent these findings apply to the broader *KCNQ2*-DEE population, whether any potential neurodevelopmental benefit of SCB is independent of seizure control, and, importantly, whether earlier treatment initiation leads to greater effectiveness in terms of both seizure control and improving neurodevelopment.

In this study, we leverage a large, deeply phenotyped cohort of 282 individuals with *KCNQ2* variants to retrospectively evaluate the effectiveness of ASMs, including SCBs, on seizure control. We further investigated whether early SCB initiation in individuals carrying a variant predicted to lead to *KCNQ2*-DEE was associated with improved neurodevelopmental outcomes and explored whether this relationship might be independent of seizure control. Our findings support that SCBs are the most effective treatment for seizure management in both SeL(F)NE and LOF *KCNQ2*-DEE, and that starting SCB therapy during the first month of life is linked to more favourable neurodevelopmental trajectories among individuals with LOF *KCNQ2*-DEE variants. These beneficial neurodevelopmental effects appear to be primarily mediated by early seizure freedom. However, the high collinearity between early SCB initiation and early seizure control complicates efforts to disentangle any potential direct disease-modifying effects of SCBs from their impact on seizure cessation, a challenge commonly observed in DEEs with onset in the first few months of life.

## Materials and methods

### Study design and participants

We conducted a multicentre retrospective study collecting detailed information on individuals with pathogenic or likely pathogenic *KCNQ2* variants. Participants were recruited through an international network of epilepsy and genetic centres, several (inter)national *KCNQ2* patient organisations, the Network for Therapies in Rare Epilepsies (NETRE), and via the European Reference Network (ERN) ERN-EpiCare Genetic Platform. French patient data were collected via the French registry *IMPROVE*: Registry of *KCNQ2*-related Epilepsy. Participants were prospectively enrolled between February 2021 and May 2025 as part of the study. Genetic and clinical data were collected using a standardised data collection sheet distributed internationally among paediatric and adult neurologists. For most participants, clinical data were collected retrospectively, with some individuals also followed prospectively. Only individuals with a minimum follow-up age of three years were included to ensure reliable neurodevelopmental outcome assessment.

### *KCNQ2* variant classification

Information on each *KCNQ2* variant, including mode of inheritance (i.e. inherited or *de novo*), was collected. Variant nomenclature was based on the *KCNQ2* transcript NM_172107.4. Variants were classified according to the American College of Medical Genetics and Genomics/Association for Molecular Pathology (ACMG/AMP) guidelines.^24^ Only individuals with (likely) pathogenic variants were included. Individuals with known mosaicism for *KCNQ2* were excluded, as mosaic status is known to influence the phenotype.^25^

Participants were stratified into three groups based on the predicted functional effect of the *KCNQ2* variant: (1) severe LOF variants associated with DEE (LOF-DEE), (2) milder LOF variants associated with SeL(F)NE (LOF-SeL(F)NE), and (3) GOF variants. Variant classification was performed according to criteria established by the RIKEE curation team of the international *KCNQ2* patient registry (www.rikee.org), integrating aggregated phenotype data from published cases and registry entries, computational predictions, functional assay results, familial segregation, and inheritance patterns (Table S1). Publicly available RIKEE classifications (https://www.rikee.org/KCNQ2-variant-database) were updated using the most recent data. Individuals carrying a *KCNQ2* variant classified as of uncertain severity were excluded. Although the RIKEE classification system does not differentiate between DEE due to LOF or GOF variants, we incorporated a GOF subgroup, informed by published literature describing variants with GOF effects, as this is recognized as a distinct clinical entity with unique phenotypic and possible therapeutic implications.^11,12,26^

### Clinical data collection

Clinical data collection included epilepsy history, ASM use and effect, as well as neurodevelopmental milestones and outcomes. To obtain a complete epilepsy history, detailed information was collected on age at seizure onset, initial seizure semiology, and seizure evolution over time. Seizures were classified following the latest ILAE guidelines.^27^ Detailed seizure descriptions were used when available; otherwise, the treating physician’s classification was retained. In neonates, reported generalised tonic-clonic seizures— an event considered unlikely given that neonates typically exhibit focal-onset seizures—were classified as sequential seizures according to neonatal ILAE classification.^28^ A period of seizure offset was defined as a seizure-free interval lasting at least 12 months, in line with the ILAE task force definitions, as this is typically longer than three times the individual’s longest inter-seizure interval.^29^ Simple febrile seizures or seizures triggered by poor adherence or abrupt ASM discontinuation, with immediate seizure freedom after reintroduction, were not considered recurrences.

Information on ASMs included all ASMs ever used and their effectiveness in seizure control. Treatment response was retrospectively classified as: highly effective for ≥90% reduction in seizure frequency; partially effective for a reduction between 50% and 90%; no effect for no change in seizure frequency; and worsening if seizure frequency increased. Reported adverse effects were also collected. Additional details were retrieved for SCBs, namely carbamazepine (CBZ), oxcarbazepine (OXC), lacosamide (LCM), lamotrigine (LTG), and phenytoin (PHT), including exact age at initiation and treatment duration. Early initiation of SCBs was defined as starting within the first month of life (neonatal period), coinciding with the typical age of seizure onset. A temporal relationship between SCB treatment and seizure freedom was considered if a seizure-free period began within two months of SCB initiation, allowing for titration to an effective dose. Individuals with less than 6 months of SCB exposure were excluded from neurodevelopmental outcome analyses to avoid including very short treatment periods unlikely to influence developmental trajectory. The 6-month cutoff was chosen pragmatically to balance the need for sufficient treatment duration with retention of an adequate sample size.

### Neurodevelopmental outcome assessment

Major neurodevelopmental milestones—age of achieving head control, independent sitting, independent walking, and first words—were recorded. Cut-offs were based on the age by which at least 95% of children achieve each milestone, according to the WHO Multicentre Growth Reference Study (MGRS)^30^ and Centres for Disease Control and Prevention (CDC) guidelines.^31^ Additional data included walking and language abilities and a general neurodevelopmental assessment at last follow-up.

Neurodevelopmental outcome was classified in line with the DSM-5 criteria^32^ and the Vineland Adaptive Behaviour Scales and Bayley Scales.^33^ The outcome was based on clinical assessment by the treating provider taking into account the level of cognitive development, adaptive functioning (daily living skills, socialisation), motor skills and language and communication. Classification levels were: normal (age-appropriate), mild (minor delays, mostly independent), moderate (noticeable delays, regular support needed), severe (major delays, dependent for most activities), and profound (extremely limited, completely dependent) impairment. When available, standardised neuropsychological assessments were used.

### Statistical analysis

Statistical analyses were performed using R (version 4.4.2). Demographic and clinical characteristics were summarised using descriptive statistics. Because the continuous variables were not normally distributed, medians with interquartile ranges (IQR) are reported. ASM effects were summarised descriptively and visualised using stacked bar plots. Categorical variables were compared using chi-square tests with Monte Carlo simulation (*B =* 10,000) where appropriate. Group differences in continuous variables were assessed using the Kruskal-Wallis rank sum test due to non-normal distributions. Associations between continuous variables were evaluated using Pearson correlation coefficients. A linear regression analysis was also performed, and results were visualised using scatter plots with fitted regression lines to illustrate the correlation. Analyses were stratified by variant subgroup (LOF-DEE, SeL(F)NE, GOF) as appropriate. Statistical significance was set at *P* < 0.05.

### Ethics

Informed consent for participation and publication was obtained from all parents or legal guardians according to the Declaration of Helsinki. The study was approved by the Human Research Ethics Committees of the University Hospital of Antwerp (Belgium, number: 20/50/683) and the Committee for the Protection of Persons (Comité de Protection des Personnes Sud-Est III, France, IDRCB: 2020-A01363-36). Data are reported in line with the Strengthening Reporting of Observational Studies in Epidemiology (STROBE) statement.

## Results

Data about 282 individuals from 21 different countries was collected. After evaluation of the genetic variant, 276 individuals were retained, and six were excluded (one variant of uncertain significance and five individuals with mosaic pathogenic variants). 233/276 (84%) individuals were aged at least 36 months at last follow-up. Among these 233 individuals, 112 carried a variant predicted to cause SeL(F)NE, 109 carried variants associated with LOF-DEE, and nine carried GOF variants. Three *KCNQ2* variants could not be classified in one of these groups based on available data and were excluded from further analyses.

### Genetic and clinical characteristics of the cohort

The genetic and clinical characteristics of the 230 individuals included in this study are summarised in Table 1. Variants in the SeL(F)NE group were equally divided between missense and truncating types and distributed throughout the Kv7.2 subunit, whereas variants in the LOF-DEE and GOF groups were almost exclusively missense and predominantly located within transmembrane regions, notably helices S4, S5, and S6, including the pore loop. Inheritance patterns also differed by group: most SeL(F)NE variants were inherited, while variants in the LOF-DEE and GOF groups occurred predominantly *de novo*. In three of the six individuals with an inherited LOF-DEE variant, the variant was proven to be inherited from a mosaic parent.

**Table 1:**
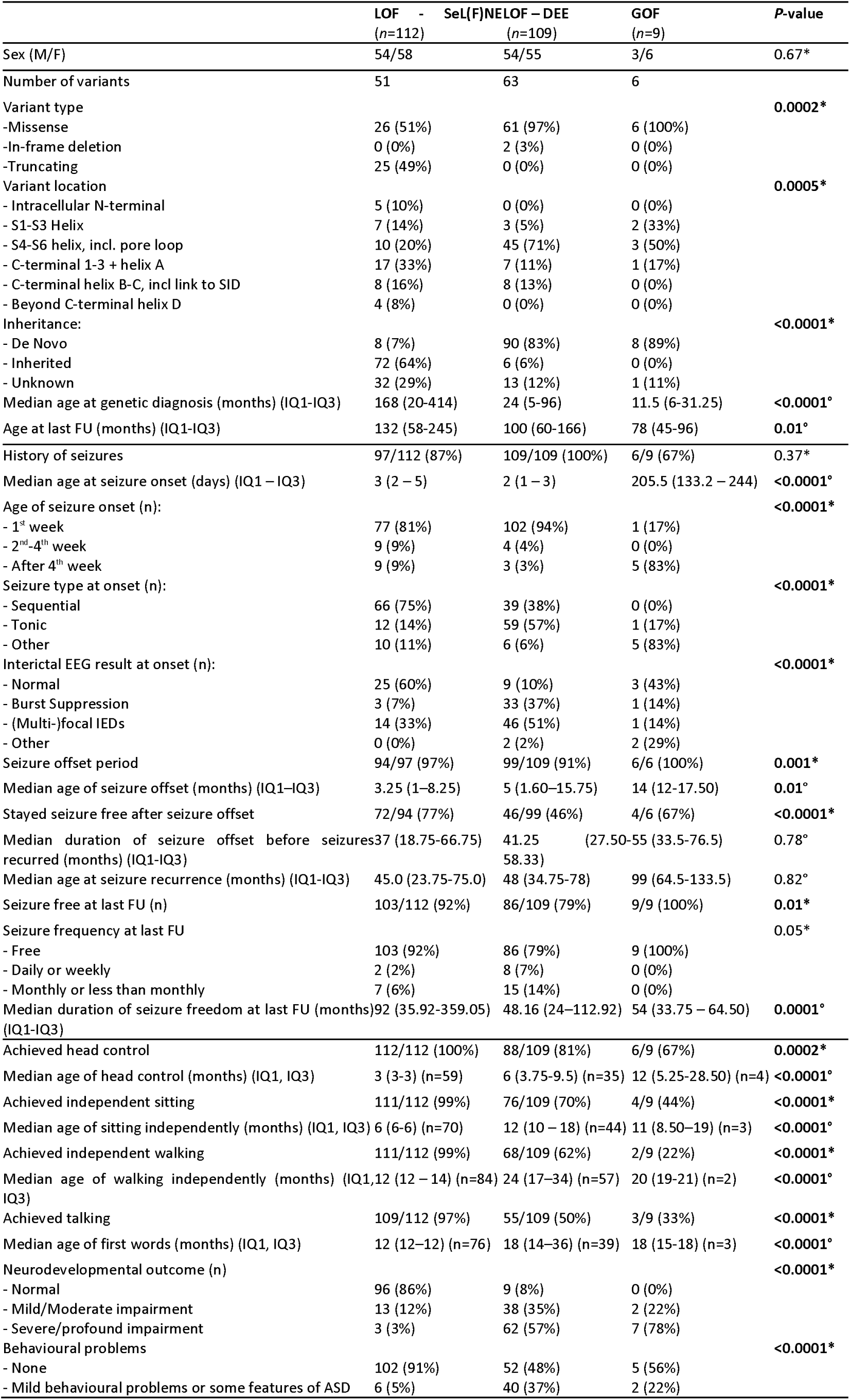
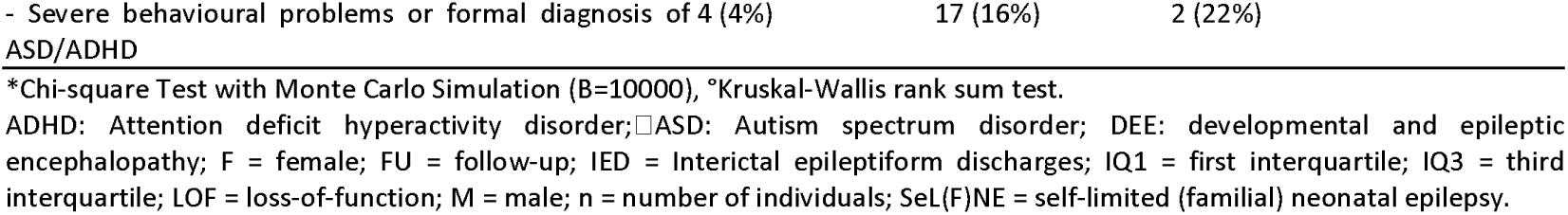
Genetic and clinical characteristics of individuals with (likely) pathogenic KCNQ2 variants stratified by predicted functional subgroup.

Distinct epilepsy profiles were observed across groups: all individuals in the LOF-DEE variant group (100%) and most in the SeL(F)NE variant group (87%) had a history of seizures, compared to 67% in the variant GOF group. The few individuals in the SeL(F)NE variant group without seizures (13%) were all asymptomatic carriers. The median age at seizure onset occurred in the first week of life for both SeL(F)NE and LOF-DEE variant groups (3 and 2 days, respectively), whereas seizure onset in the GOF variant group occurred later (median 206 days). Age at seizure onset was not adjusted for gestational age, as this data was not available for all participants; only 23 individuals (eight SeL(F)NE, fifteen LOF-DEE) were reported as premature, of whom two had seizure onset after the neonatal period. The exact gestational age at birth for these two patients was not available. Seizure types at onset also varied: sequential seizures were most common in case of SeL(F)NE variants, tonic seizures predominated for LOF-DEE variants, and a range of seizure types at onset was observed for GOF variants, including generalised tonic-clonic, generalised myoclonic, epileptic spasms, focal with impaired consciousness, and focal tonic. Seizure offset was achieved by the majority in all groups. Neurodevelopmental milestones were most frequently attained in the SeL(F)NE variant group, while median ages of acquisition were later in the LOF-DEE and GOF variant groups if these milestones were achieved.

### Effects of antiseizure medications on seizures in LOF-DEE and SeL(F)NE cohorts

ASM treatment history and their reported effects on seizures, combining all treatment initiation periods, were analysed for all individuals who experienced seizures with LOF variants (97 SeL(F)NE, 109 LOF-DEE). ASM history was unavailable for two individuals, both from the SeL(F)NE group. Due to the limited number of individuals with epilepsy (*n* = 6) in the GOF subgroup, this group was excluded from the analysis.

Over the disease course, 186 documented treatment effects from 19 ASMs were recorded in the SeL(F)NE cohort, versus 470 effects from 22 ASMs in the LOF-DEE cohort. The median number of ASMs used per individual was significantly lower in the SeL(F)NE group (2; interquartile range [IQR] 1-3) compared to the LOF-DEE group (4; IQR 3-6) (*P* < 0.0001, Kruskal-Wallis rank sum test). The effects of all administered ASMs on seizures in both cohorts are detailed in Supplementary Fig. 1, while Fig.1 highlights the treatment responses to the most frequently used ASMs in these groups.

**Figure 1:**
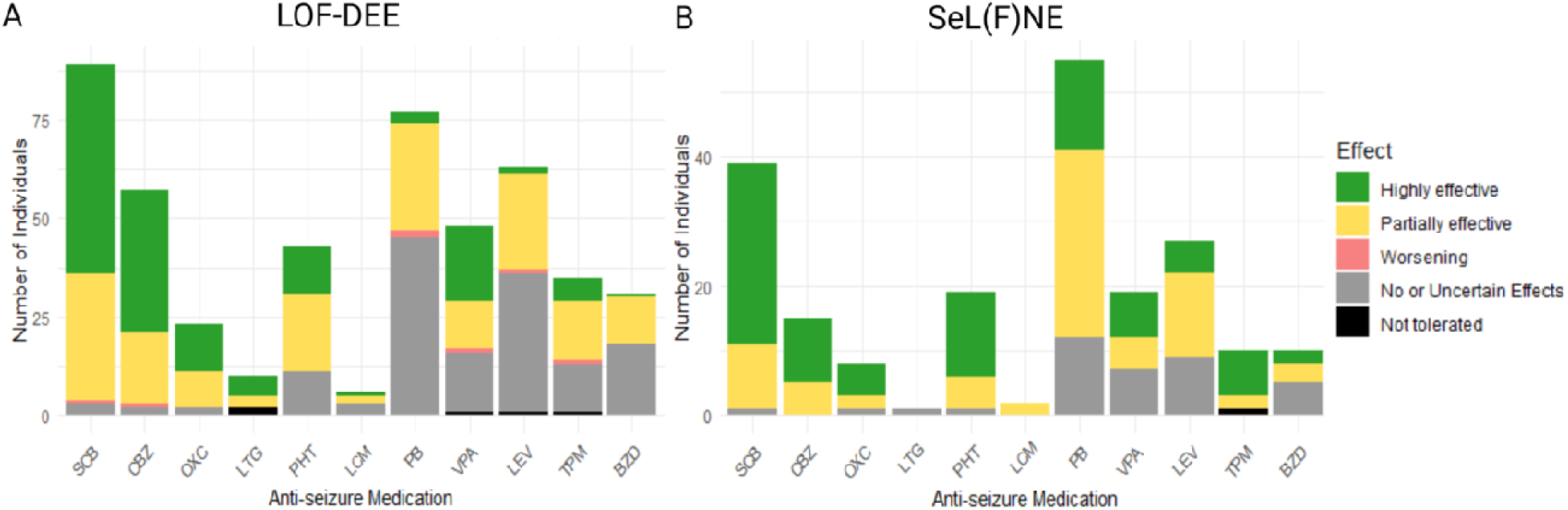
Effects of most frequently used antiseizure medications (ASMs) on seizures in LOF-DEE and SeL(F)NE cohorts. The figure shows reported treatment effects of ASMs among individuals with (likely) pathogenic *KCNQ2* variants predicted to lead to **(A)** loss-of-function developmental and epileptic encephalopathy (LOF-DEE) and **(B)** self-limited familial neonatal epilepsy (SeL(F)NE). Effects are categorised as highly effective (>90% seizure reduction), partially effective (reduction in seizure frequency between 50% and 90%), worsening (increase in seizure frequency), no or uncertain effects, and not tolerated. The stacked bar plots summarise the number of individuals experiencing each effect per ASM. For the category labelled ‘SCB’ (sodium channel blockers), the data represents a composite effect reflecting the best reported response among the individual SCB ASMs (CBZ, OXC, LTG, PHT, and LCM) administered. For example, if an individual experienced a partial effect with phenytoin (PHT) but a highly effective response with carbamazepine (CBZ), the ‘SCB’ category records the highest effect (i.e., highly effective) for that individual. This approach consolidates the overlapping mechanisms of these drugs in the analysis. Similarly, the ‘BZD’ (benzodiazepines) category summarises the combined effects of all benzodiazepines used by each individual, reflecting the overall BZD treatment effect regardless of specific drug. ASMs = antiseizure medications; BZD = benzodiazepines; CBZ = carbamazepine; LCM = lacosamide; LEV = levetiracetam; LOF-DEE = loss-of-function developmental and epileptic encephalopathy; LTG = lamotrigine; OXC = oxcarbazepine; PB = phenobarbital; PHT = phenytoin; SeL(F)NE = self-limited (familial) neonatal epilepsy; SCB = sodium channel blockers; TPM = topiramate; VPA = valproate.

A significantly higher proportion of highly effective treatment responses was observed in the SeL(F)NE group (68/186, 37%) compared to the LOF-DEE group (101/470, 21%; *P* < 0.0001, Chi-square test). However, in both cohorts, SCBs, including CBZ, OXC, PHT, LTG and LCM, were most frequently reported as highly effective, accounting for 28/68 (41%) responses in the SeL(F)NE group and 66/101 (65%) in the LOF-DEE group, observed among 28 and 53 individuals, respectively (Fig. 1). Among SCBs, CBZ, PHT, and OXC were the most commonly used. In the LOF-DEE group, CBZ and OXC were more commonly reported as highly effective (36/57, 63%; 12/23, 52%, respectively) compared to PHT (12/43, 28%; *P* = 0.002, Chi-square test). Within the SeL(F)NE group, effectiveness rates of these three SCBs were comparable (CBZ: 10/15, 67%; OXC: 5/8, 63%; PHT: 13/19, 68%; *P* = 1, Chi-square test with Monte Carlo simulation). Due to limited exposure numbers, conclusions regarding LTG and LCM could not be drawn.

Other ASMs such as valproate (VPA), phenobarbital (PB), topiramate (TPM), and levetiracetam (LEV) were reported to be effective in subsets of both SeL(F)NE and LOF-DEE cases. Notably, in the LOF-DEE group, VPA showed second-best rate of high efficacy (19/48, 40%) which was significantly greater than those for PB (3/77, 4%) and LEV (2/63, 3%) (*P* < 0.0001, Chi-square test with Monte Carlo simulation).

### Detailed analysis of SCB initiation and its association with seizure freedom

Given that SCBs were reported as the most effective ASMs, we conducted a more detailed analysis focusing on the timing of SCB initiation and its relationship to seizure offset. Among individuals in the SeL(F)NE and LOF-DEE groups, 34/97 (35%) and 85/109 (78%) individuals, respectively, had received at least one SCB. The timing of SCB initiation was unavailable for two individuals (one SeL(F)NE and one LOF-DEE). The median age at SCB initiation did not differ significantly (SeL(F)NE: median 91 days, IQR 23–370; LOF-DEE: median 39 days, IQR 10–365; *P* = 0.199, Kruskal–Wallis rank sum test).

Within the SeL(F)NE group, 10/33 individuals (30%) initiated SCBs within the first month of life (the seizure onset period for >90% of this cohort), compared to 41/84 (49%) in the LOF-DEE group. In both groups, earlier SCB initiation was associated with substantially earlier seizure offset. Specifically, the median age at seizure offset was 1.6 months (IQR 0.75–9; n = 41) in the LOF-DEE group and 1 month (IQR 1–2.75; n = 10) in the SeL(F)NE group among those who started SCBs in the first month of life. In contrast, seizure offset occurred at a median of 5 months (IQR 2.5–11; n = 43) in later starters in the LOF-DEE group and at 4 months (IQR 3–26; n = 23) in the SeL(F)NE group, while those who never received SCBs had the latest offsets (LOF-DEE: 15 months, IQR 6–39; n = 22; SeL(F)NE: 3 months, QR: 1-6.75; n=56). These differences were statistically significant (LOF-DEE: *P* = 0.0008; SeL(F)NE: *P* = 0.01; Kruskal–Wallis test).

Among individuals who experienced a period of seizure offset, a significantly smaller proportion of the SeL(F)NE group were taking SCBs at the time of seizure offset compared to the LOF-DEE group (29/88, 33% vs. 56/96, 58%; *P* = 0.0015, Chi-square test). For those who did achieve seizure offset while on SCBs, a temporal relationship was observed in 17/29 (59%) in the SeL(F)NE group and 41/56 (73%) in the LOF-DEE group. In addition, we observed strong positive correlations between the timing of SCB initiation and age at subsequent seizure offset in both groups (Fig. 2).

**Figure 2:**
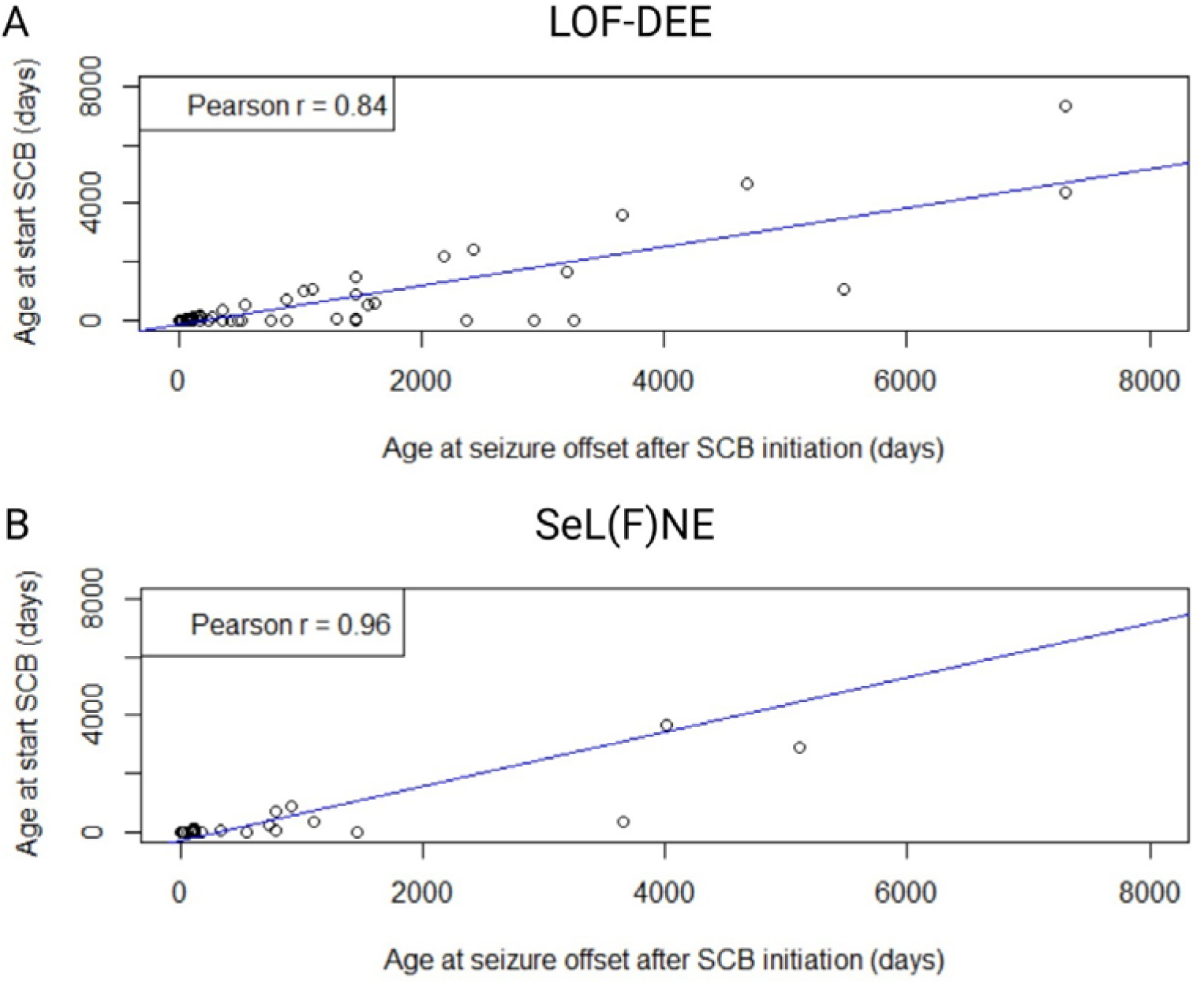
Correlation between age at sodium channel blocker (SCB) initiation and age at subsequent seizure offset in the (A) developmental and epileptic encephalopathy (DEE) and the (B) self-limited (familial) neonatal epilepsy (SeL(F)NE) cohorts. Scatter plots depicting the relationship between age at SCB initiation and age at subsequent seizure freedom. Panel **(A)** shows data from the DEE cohort, revealing a strong positive correlation with a significant linear regression (Pearson r = 0.84, *P* < 0.0001). The linear regression model for this subset showed a significant slope (β = 0.66, SE = 0.050, t-value=13.12; *P* <0.001), explaining 70.22% of the variance. Panel **(B)** shows data from the SeL(F)NE cohort, revealing a strong positive correlation with a significant linear regression (Pearson r = 0.96, *P* < 0.0001). The linear regression model for this subset showed a significant slope (β = 0.93, SE = 0.049, t-value=18.96; *P* <0.001), explaining 93% of the variance. Blue regression lines indicate the fitted linear models for each subset.

Moreover, when examining the timing of SCB initiation in relationship with seizure outcomes, no statistically significant difference was observed between treatments initiated within the first month or first year of life and those introduced later, indicating that SCBs remain effective for seizures even when administered at more advanced stages (Supplementary Fig. 2).

### More frequent and earlier use of SCBs in LOF *KCNQ2*-related disorders

Since SCBs were first recommended as first-line therapy for neonatal seizures in *KCNQ2*-related disorders in 2015,^19^ we investigated whether their use has become more frequent and initiated earlier in younger individuals. Cases with *KCNQ2* GOF variants were excluded from this analysis, as they typically do not present with neonatal seizures. The median year of birth for individuals who used SCBs was 2013 (Q1: 2008, Q3: 2018, *n*=119), compared to 2008 (Q1: 1988, Q3: 2015, *n*=78) for those who never received them. Among individuals born in 2015 or earlier, 65 out of 124 (52%) received SCBs at some point during their life, compared to 54 out of 73 (74%) of those born after 2015 (*P* = 0.005, Chi-squared test). Among individuals born after 2015 who received SCBs, 18 out of 54 (33%) started treatment within the first month of life, compared to 6 out of 65 (9%) of those born in 2015 or earlier. A significant negative correlation was also observed between year of birth and age at SCB initiation, indicating that children born more recently started SCB therapy at younger ages (Supplementary Fig. 3).

### Early SCB use is associated with better developmental milestone outcomes

Given that most individuals carrying SeL(F)NE variants exhibit generally normal neurodevelopmental outcomes, we focused our investigation of (early) SCB treatment effects on the subgroup with LOF-DEE variants (*n* = 106; data unavailable for three). After excluding seven individuals with less than six months of SCB exposure and three with incomplete neurodevelopmental data, the cohort was divided into three groups based on timing of SCB initiation: early (within the first month of life, *n* = 36), late (after the first month, *n* = 38), and never treated with SCBs (*n* = 22). Median duration of SCB treatment was comparable between early and late starters (41 and 42 months, respectively), though with wide variability (IQR 16.8-67.5 and IQR 24.2-89.2, respectively).

Individuals who initiated SCB therapy early showed significantly higher rates of achieving key milestones—including head control (particularly before 4 months), independent sitting (particularly before 9 months), independent walking, and talking—compared to those with later or no SCB exposure (Table 2, Fig. 3).

**Figure 3:**
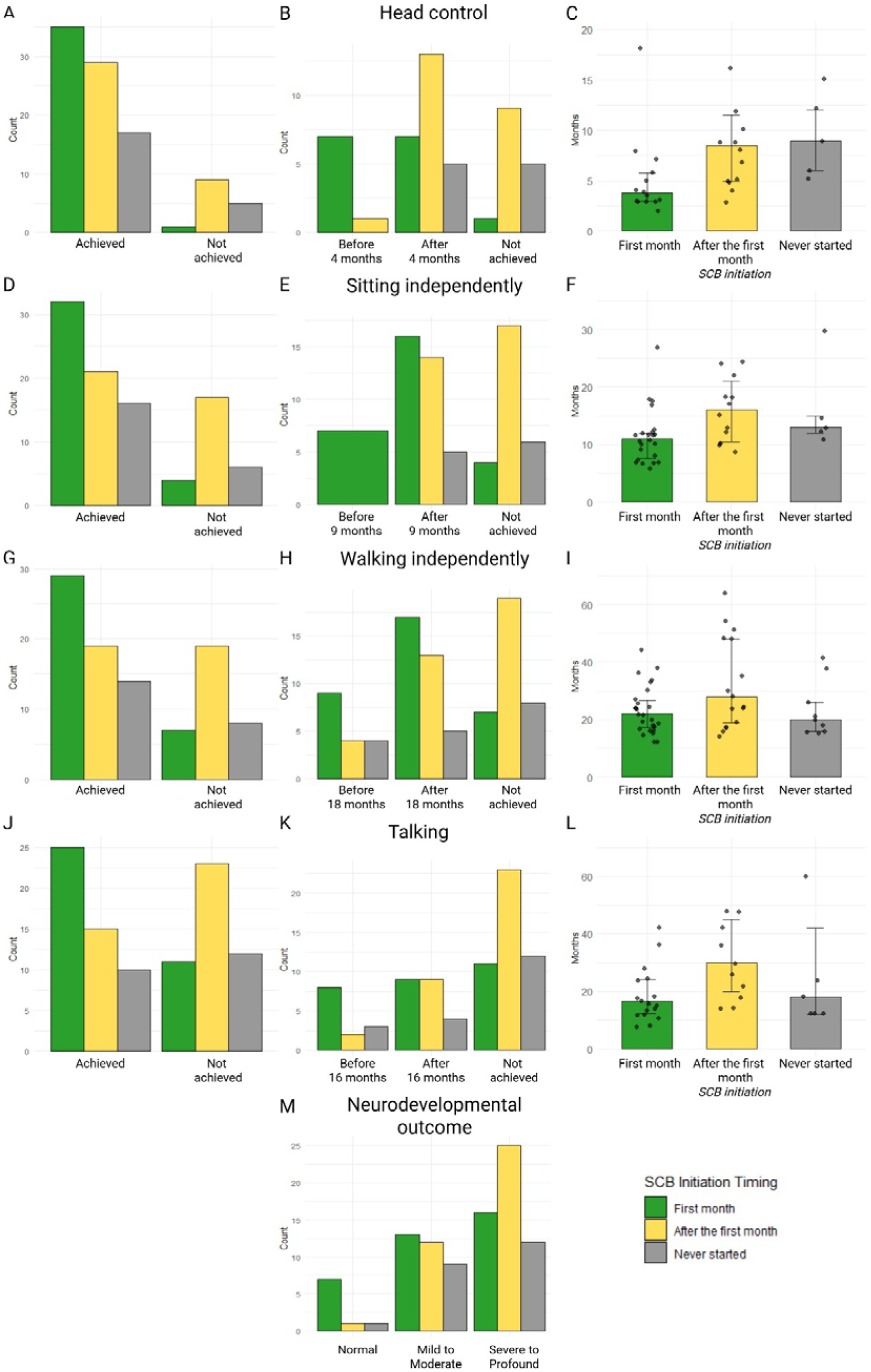
Neurodevelopmental milestones and outcomes by timing of sodium channel blocker initiation in individuals with *KCNQ2*-LOF DEE variants. Panels **A–L** present the attainment and timing of key neurodevelopmental milestones in individuals according to sodium channel blocker (SCB) initiation group: within the first month of life (green), after the first month (yellow), and never started SCBs (grey). For each milestone (head control, independent sitting, independent walking, and first words/talking), three panels are shown: **(A, D, G, J)** proportions of individuals who achieved the milestone versus those who did not, **(B, E, H, K, M)** distribution across specific age categories for milestone attainment, and **(C, F, I, L)** median age of milestone achievement with interquartile ranges (IQR) and individual data points overlaid. Panel **M** summarises overall neurodevelopmental outcome as classified into normal, mild/moderate, or severe/profound impairment categories per treatment group. Bar heights, colours, and dots correspond to the data presented in the accompanying summary Table 2.

**Table 2:**
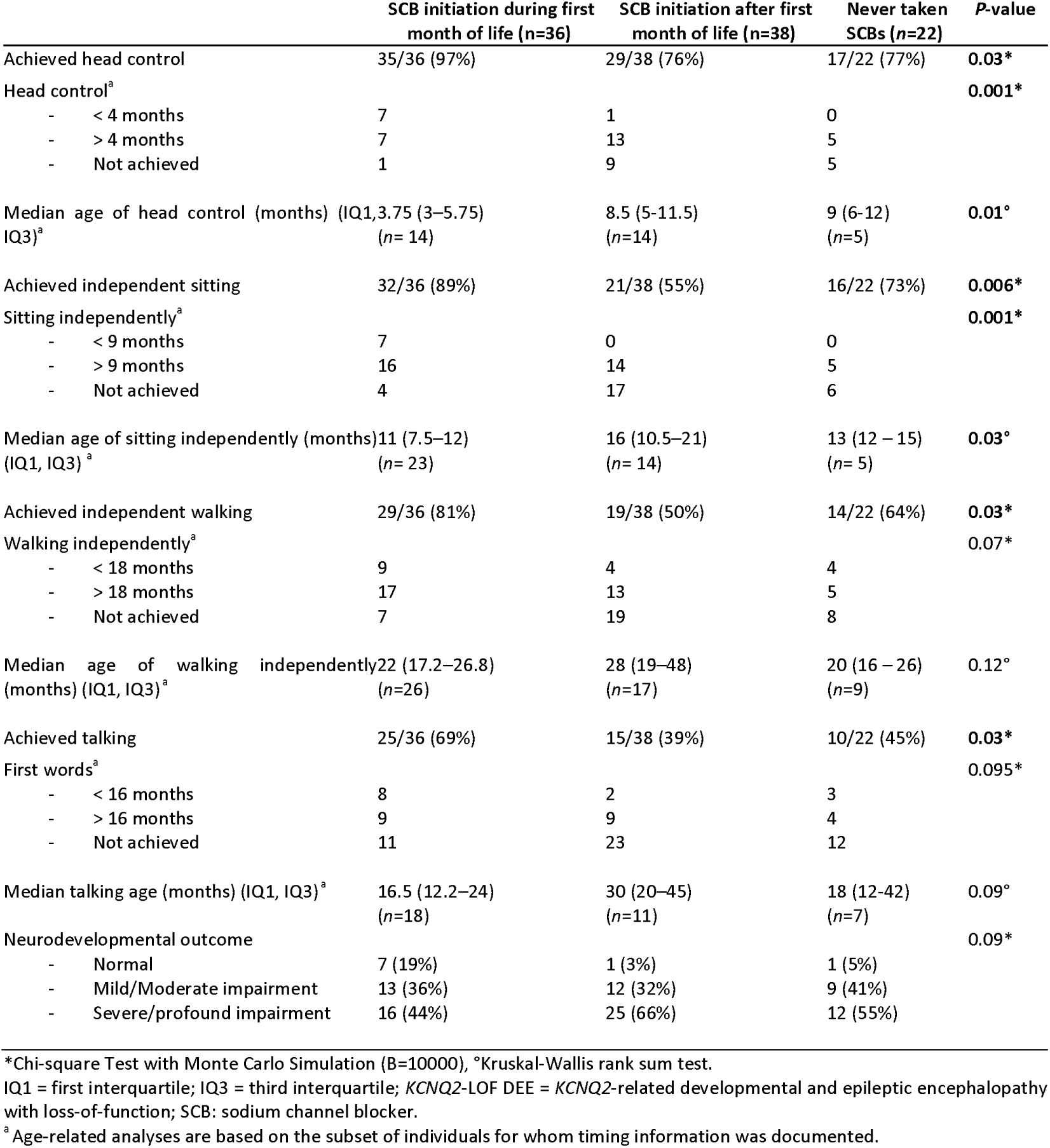
Neurodevelopmental milestones and outcomes stratified by timing of sodium channel blocker initiation in individuals with KCNQ2-LOF DEE variants.

|  | SCB initiation during first month of life (n=36) | SCB initiation after first month of life (n=38) | Never taken SCBs (n=22) | P-value |
| --- | --- | --- | --- | --- |
| Achieved head control | 35/36 (97%) | 29/38 (76%) | 17/22 (77%) | **0.03\*** |
| Head control<sup>a</sup> |  |  |  | **0.001\*** |
| - < 4 months | 7 | 1 | 0 |  |
| - > 4 months | 7 | 13 | 5 |  |
| - Not achieved | 1 | 9 | 5 |  |
| Median age of head control (months) (IQR)<sup>a</sup> | 3.75 (3–5.75)<br>(n=14) | 8.5 (5–11.5)<br>(n=14) | 9 (6–12)<br>(n=5) | **0.01°** |
| Achieved independent sitting | 32/36 (89%) | 21/38 (55%) | 16/22 (73%) | **0.006\*** |
| Sitting independently<sup>a</sup> |  |  |  | **0.001\*** |
| - < 9 months | 7 | 0 | 0 |  |
| - > 9 months | 16 | 14 | 5 |  |
| - Not achieved | 4 | 17 | 6 |  |
| Median age of sitting independently (months) (IQR)<sup>a</sup> | 11 (7.5–12)<br>(n=23) | 16 (10.5–21)<br>(n=14) | 13 (12–15)<br>(n=5) | **0.03°** |
| Achieved independent walking | 29/36 (81%) | 19/38 (50%) | 14/22 (64%) | **0.03\*** |
| Walking independently<sup>a</sup> |  |  |  | **0.07\*** |
| - < 18 months | 9 | 4 | 4 |  |
| - > 18 months | 17 | 13 | 5 |  |
| - Not achieved | 7 | 19 | 8 |  |
| Median age of walking independently (months) (IQR)<sup>a</sup> | 22 (17.2–26.8)<br>(n=26) | 28 (19–48)<br>(n=17) | 20 (16–26)<br>(n=9) | **0.12°** |
| Achieved talking | 25/36 (69%) | 15/38 (39%) | 10/22 (45%) | **0.03\*** |
| First words<sup>a</sup> |  |  |  | **0.095\*** |
| - < 16 months | 8 | 2 | 3 |  |
| - > 16 months | 9 | 9 | 4 |  |
| - Not achieved | 11 | 23 | 12 |  |
| Median talking age (months) (IQR)<sup>a</sup> | 16.5 (12.2–24)<br>(n=18) | 30 (20–45)<br>(n=11) | 18 (12–42)<br>(n=7) | **0.09°** |
| Neurodevelopmental outcome |  |  |  | **0.09\*** |
| - Normal | 7 (19%) | 1 (3%) | 1 (5%) |  |
| - Mild/Moderate impairment | 13 (36%) | 12 (32%) | 9 (41%) |  |
| - Severe/profound impairment | 16 (44%) | 25 (66%) | 12 (55%) |  |

\*Chi-square Test with Monte Carlo Simulation (B=10000), °Kruskal-Wallis rank sum test.
IQR = first interquartile; IQR = third interquartile; *KCNQ2*-LOF DEE = *KCNQ2*-related developmental and epileptic encephalopathy with loss-of-function; SCB: sodium channel blocker.
<sup>a</sup> Age-related analyses are based on the subset of individuals for whom timing information was documented.

Additional analyses were performed using alternative age cut-offs for SCB initiation, as summarised in Supplementary Tables 2–4. When these alternative groupings were applied, many of the statistically significant differences in neurodevelopmental milestone acquisition and outcome observed with the original cut-off (first month vs later vs never) were attenuated or no longer reached significance.

### Early seizure freedom is associated with improved neurodevelopmental outcomes

Next, we analysed neurodevelopmental outcomes in relation to the timing of seizure offset. We first examined age at seizure offset as a continuous variable against achievement of major developmental milestones and overall outcome. Among the 99 individuals who experienced a period of seizure offset in the *KCNQ2* LOF-DEE subgroup, earlier acquisition of developmental milestones such as head control, sitting, talking, and favourable neurodevelopmental outcome was associated with younger median age at seizure offset (Fig. 4). For the achievement of walking, similar trends were observed, although these did not reach statistical significance (*P* = 0.14).

**Figure 4:**
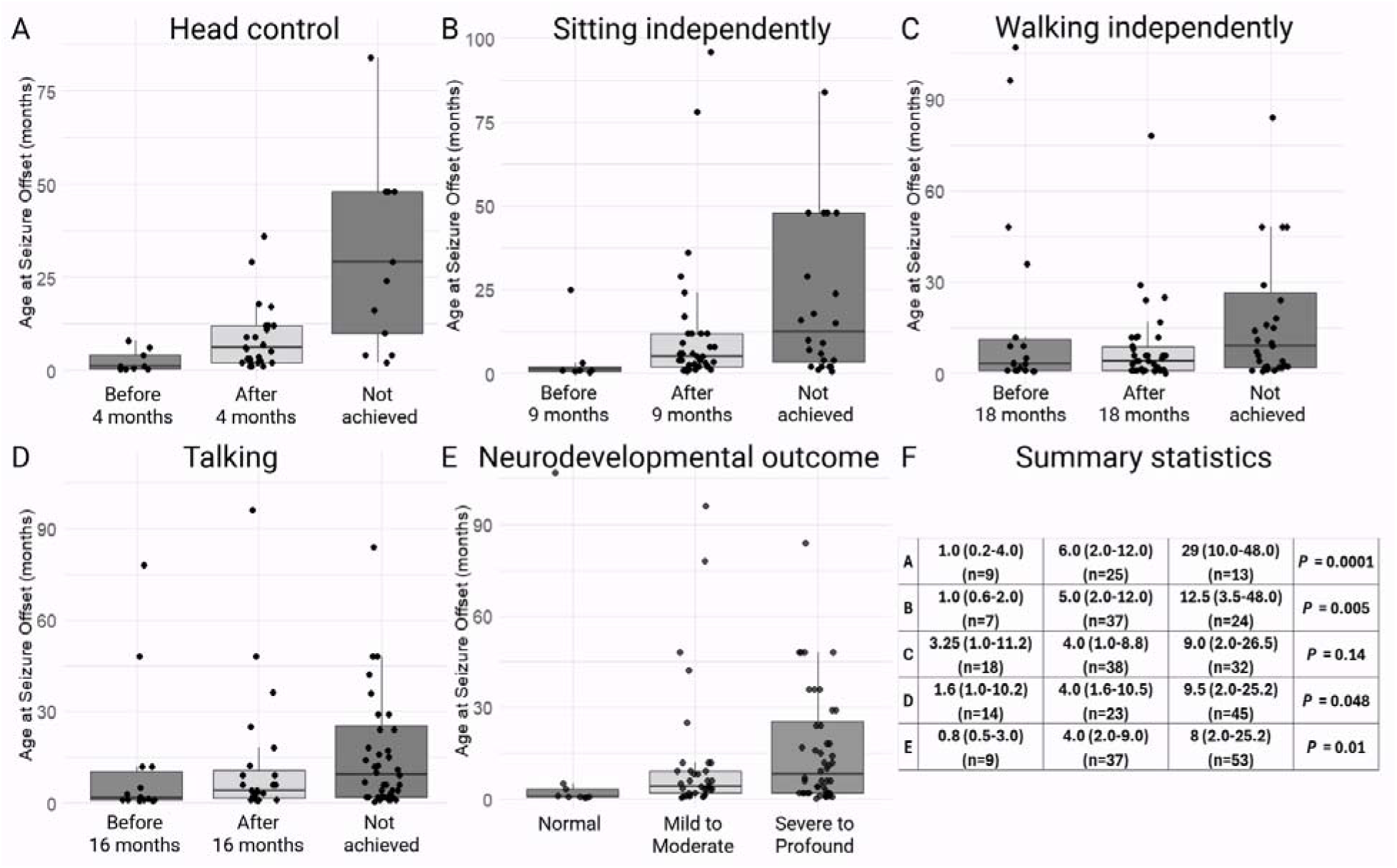
Timing of seizure offset in relation to neurodevelopmental milestones and outcome. Bar plots (Panels **A–E**) and summary table (Panel **F**) show the distribution of age at seizure offset across categories of major developmental milestones (head control, independent sitting, walking, talking) and neurodevelopmental outcome in individuals with a *KCNQ2*-DEE variant who experienced a period of seizure freedom (*n* = 99). For each outcome, age at seizure offset is reported as median (IQ1–IQ3) for each category (e.g., <4 months, >4 months, not achieved for head control). Significant associations were observed for head control, independent sitting, talking, and overall neurodevelopmental outcome (Kruskal-Wallis test, all *P* ≤ 0.005), while trends for walking did not reach significance. Earlier developmental milestone attainment and favourable neurodevelopmental outcomes are correlated with younger ages at seizure offset.

Because the timing of seizure freedom appeared to strongly influence developmental outcomes, we next evaluated whether specific postnatal age cut-offs for seizure offset might define clinically relevant ‘windows of opportunity’ for improved prognosis. Guided by both the natural ages for milestone attainment and observed patterns from the data, cut-offs of 1 (Table 3), 2, 3, 4, 6, 8, and 12 months (Supplementary Tables 5–13) were selected for focused analysis. In these analyses, individuals who never became seizure free were included as a distinct group to capture their unique developmental trajectory.

**Table 3:**
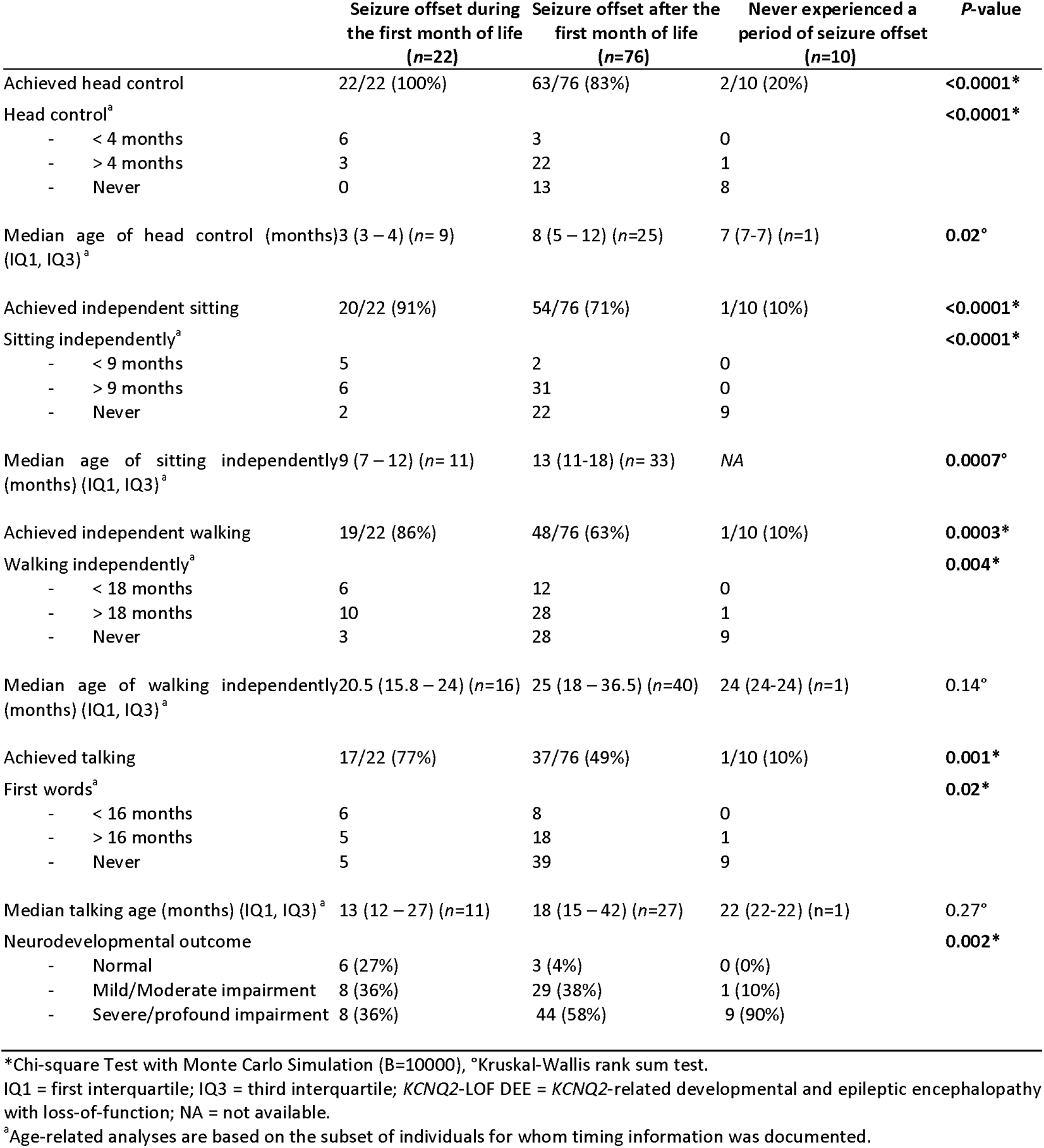
Neurodevelopmental milestones and outcomes stratified by timing of seizure offset in individuals with KCNQ2-LOF DEE variants.

The clearest distinction in outcome was observed with a 1-month threshold (Table 3), supporting this period as particularly critical for neurodevelopment in *KCNQ2*-LOF DEE, with later or never offset associated with lower rates of achieving key gross motor milestones and more severe delays. Broader cut-offs (Supplementary Tables 5–13) generally supported these findings, although some statistical significances were attenuated or lost.

### Does early SCB use impact neurodevelopment outcomes beyond its effect on seizures?

Our results show that both SCB initiation in the first month as well as earlier seizure freedom are associated with significantly better attainment of developmental milestones and neurodevelopmental outcomes. Therefore, we evaluated whether early SCB use influences neurodevelopmental outcomes independently of early seizure control. Because 18/22 (82%) of individuals that became seizure free during the first month of life also initiated SCB during the neonatal period, a direct comparison between those who achieved seizure freedom with SCBs versus those who achieved this with other ASMs was not feasible. Therefore, we stratified the data by seizure freedom timing within the subgroup of individuals who started SCB in the first month of life (n=41, Table 4).

**Table 4:**
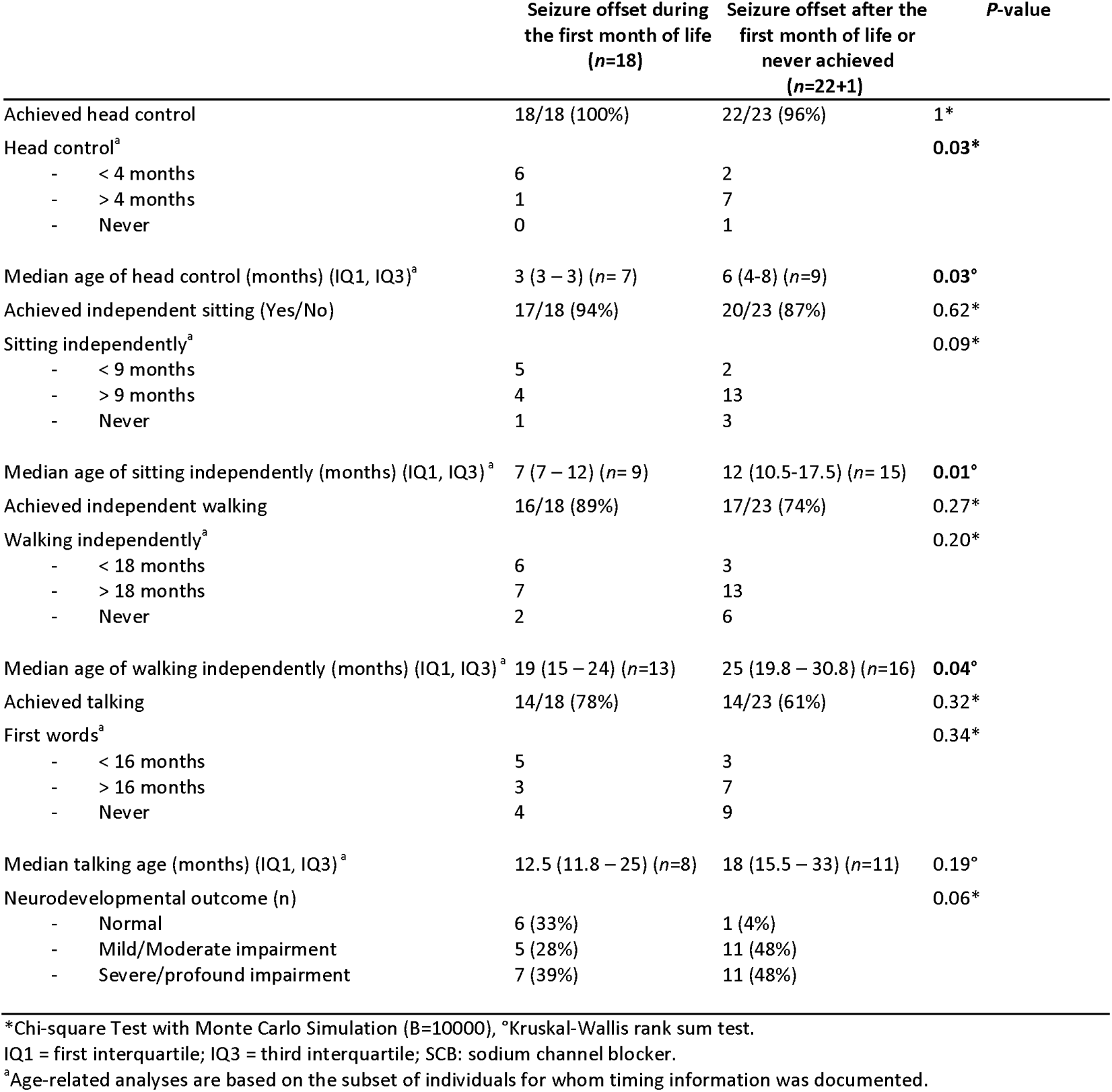
Neurodevelopmental milestones and outcomes stratified by timing of seizure offset in individuals with SCB initiation in the first month of life.

Within the subgroup that initiated SCBs in the first month of life, individuals who achieved seizure freedom during this period achieved head control, independent sitting, and walking at significantly younger ages compared to those with later or no seizure freedom (Table 4). In addition, we observed a higher proportion of individuals with normal neurodevelopmental outcome among those who achieved seizure freedom in the first month, but these differences did not reach statistical significance (*P* = 0.06). While early initiation of SCBs correlates with earlier seizure freedom, achieving seizure offset early appears to be the key determinant of better developmental outcomes, suggesting that seizure control itself, rather than other variables possibly influenced by treatment timing, is the most critical factor for neurodevelopmental prognosis.

### Influence of SCB use in individuals with recurrent *KCNQ2* LOF-DEE variants

Next, we assessed the influence of early SCB use and seizure offset in a subset of individuals carrying the same pathogenic *KCNQ2* variant (Supplementary Table 14). This analysis revealed that early seizure freedom and/or early SCB initiation does not guarantee a favourable neurodevelopmental trajectory, and a considerable phenotypic variability remains even among individuals with the same variant.

## Discussion

This multicentre international retrospective study provides a large systematically collected dataset on individuals with (likely) pathogenic *KCNQ2* variants, enabling the evaluation of treatment effects on seizures as well as neurodevelopment across individuals carrying predicted LOF SeL(F)NE and LOF-DEE variants. Our findings highlight that SCBs are the most effective ASM for individuals with LOF variants, and that early cessation of seizures, most easily achieved by SCB treatment, improves neurodevelopmental outcomes in KCNQ2 LOF-DEE.

### Epilepsy course and treatment response

In both LOF DEE and SeL(F)NE groups, seizures typically began in the neonatal period, and most individuals eventually experienced seizure remission, though the timing and likelihood of recurrence differed among patients. In line with previous retrospective series reporting more difficult to treat seizures in the LOF-DEE group,^8,9,19^ we found that seizure offset occurred sooner with lower recurrence rates, and a greater proportion of seizure-free individuals at last follow-up in the SeL(F)NE compared to LOF-DEE. This was further supported by fewer ASMs used per individual and a higher proportion of highly effective ASMs in the SeL(F)NE group, likely reflecting its self-limiting course.

Individuals were exposed to a wide range of ASMs in both SeL(F)NE and LOF-DEE cohorts, underscoring the ongoing challenge of seizure control, particularly in the LOF-DEE group. In the LOF-DEE group, VPA emerged as the second most effective ASM after SCBs, whereas in the SeL(F)NE group, PB was most frequently reported as highly effective following SCBs. As previously reported, seizures in individuals with GOF variants typically presented later in infancy and were rarely observed in the neonatal period.^6,11,12,34^ Remarkably, all GOF individuals achieved seizure freedom by the last follow-up, suggesting a distinct and more favourable seizure prognosis.

### Highest effect of sodium channel blockers on seizures in LOF *KCNQ2*-related disorders

SCBs were most frequently reported as highly effective ASMs, achieving seizure reduction of more than 90%, in both the LOF-DEE and SeL(F)NE groups, supporting prior case series and anecdotal reports.^18–20^ Both groups also showed a clear temporal correlation between the timing of SCB initiation and subsequent seizure offset. While high effectiveness was observed across all SCBs, CBZ and OXC were more often reported as highly effective than PHT in the LOF-DEE group. This difference was not observed in the SeL(F)NE group, likely reflecting the more favourable natural history and relative ease of gaining seizure control. The lower apparent effectiveness of PHT in LOF-DEE may be explained by challenges in titration and serum level monitoring due to its non-linear pharmacokinetics, especially in neonates and in a setting where seizures are more difficult to control.^35^ In addition, PHT carries a risk of long-term adverse effects. Data on other SCBs, including LTG or LCM, remain limited. LTG may be less suitable in acute settings due to its slow titration schedule while LCM, which is available in an intravenous formulation, is a promising alternative, especially given its use in convulsive status epilepticus. However, the current sample size is too small to assess efficacy of these ASMs. Therefore, based on the results of this study, CBZ or OXC should be considered first-line ASMs in LOF *KCNQ2*-related disorders, especially as they are readily available and comparatively inexpensive drugs.

It is encouraging to observe a significant trend toward earlier and more frequent SCB use in children born after 2015, likely reflecting increased awareness following key publications and recommendations.^19–21^ However, despite this progress, a notable proportion of children still did not receive a SCB, and among those who did, many were not started within the first month of life, highlighting room for improvement in clinical practice.

### Early SCB use is associated with improved neurodevelopmental outcomes

Our results indicate that initiation of SCB treatment within the first month of life is associated with more favourable neurodevelopmental trajectories in *KCNQ2*-LOF DEE. These results support the hypothesis that early therapeutic intervention during a critical developmental window may have a beneficial effect on cognitive outcomes. This is consistent with observations in a *KCNQ2*-DEE mouse model exhibiting seizures and cognitive defictis^36^, in which early and sustained SCB treatment not only reduced seizures but also enhanced cognitive function.^23^

Notably, when later initiation cut-offs were applied, statistically significant differences were attenuated, although trends toward developmental benefits remained. This pattern suggests a continuum of effect rather than a strict threshold, underscoring that starting SCBs after the first month may still confer meaningful clinical advantages. Thus, delayed initiation should not be discouraged, as it may still contribute positively to seizure control and neurodevelopmental outcomes.

### Timing of seizure freedom strongly influences neurodevelopmental outcome

Our analyses demonstrate that earlier seizure freedom is closely associated with more favourable neurodevelopmental trajectories in individuals with *KCNQ2*-LOF DEE. Specifically, seizure offset during the first month of life correlated significantly with earlier attainment of key developmental milestones such as head control and independent sitting, as well as with a higher likelihood of normal overall neurodevelopmental outcome. Subsequent analyses using broader age cut-offs largely supported these findings, though some associations were attenuated, underscoring the robust impact of early seizure control on developmental prognosis. These data align with prior work, demonstrating a relationship between higher seizure frequency and duration and more severe developmental outcome in other DEEs, such as Dravet syndrome and infantile epileptic spasm^37,38^ emphasising the importance of early seizure control, regardless of aetiology or treatment.

This concept has also been well illustrated in a preclinical mouse model that conditionally expressed dominant-negative Kv7.2 subunits in brain. Here, Peters *et al.* (2005) demonstrated that selective suppression of M-current had to be reversed during the first postnatal week to prevent permanent neurophysiological and behavioural alterations in mice.^39^ Prophylactic treatment with sodium-potassium-chloride cotransporter NKCC1 antagonist bumetanide in the same mouse model during the first two postnatal weeks, normalised network activity *in vivo,* and prevented structural and behavioural pathology.^40^

### Disentangling independent effects of SCBs on neurodevelopment: a challenge

In young children with Dravet syndrome, dose-dependent improvements in executive function have been observed with fenfluramine, during the early formative years of neurodevelopment, which were not consistently linked to seizure reduction.^41^ In line, it has been hypothesized that CBZ may exert a disease-modifying effect—possibly mediated by its active metabolite, epoxy-CBZ—by acting directly on the developing brain, independently of seizure control.^23^ Since our findings demonstrate that both early initiation of SCBs as well as early seizure freedom are associated with improved neurodevelopmental outcomes, we sought to disentangle the independent effects of early SCB use on neurodevelopment by performing subgroup analyses. Among individuals who started SCBs in the first month, those who became seizure-free within the first month reached key milestones such as head control, sitting, and walking significantly earlier, suggesting that the neurodevelopmental benefits of early SCB use are at least in part driven by earlier seizure freedom. However, high collinearity between early SCB initiation and early seizure control makes disentangling independent effects challenging. While a direct neuroprotective or disease-modifying effect of SCBs independent of seizure control remains a possibility, larger studies are needed to clarify this. For now, these data reinforce the timing of seizure freedom as the predominant driver of neurodevelopmental outcome in *KCNQ2*-LOF DEE.

### Variability in seizure and neurodevelopmental outcomes

In our LOF-DEE cohort, some individuals were not seizure-free despite early use of SCBs. Moreover, several individuals who achieved early seizure cessation, including those with neonatal SCB initiation, still had severe or profound neurodevelopmental impairment and failed to attain major milestones. These observations highlight that therapeutic response is multifactorial and cannot be fully explained by seizure control or treatment timing alone.

A possible explanation is that individuals with more severe *KCNQ2* variants experience refractory seizures or worse neurodevelopmental outcomes, making non-specific treatments like SCBs less effective. Nearly all carriers of p.(Arg560Trp) and p.(Ala265Thr) showed severe impairments despite early SCB treatment, whereas early treatment of p.(Ala196Val), p.(Ala265Val), or p.(Arg553Trp) carriers was associated with better neurodevelopmental outcomes. The functional and structural effects of the variant itself likely contribute to both seizure severity and overall developmental prognosis.^6,12^ These observations underscore the need for more precise, mechanism-targeted therapies, such as antisense oligonucleotides (ASOs) or other molecular correction strategies, that could directly address the underlying molecular dysfunction and not merely seizure control. In addition, the presence of genetic, such as common variants, may modulate phenotypic severity, as has been observed in other presumed monogenic epilepsies and DEE.^42,43^ Future large-scale genomic analyses may help to clarify these influences and aid in tailoring prognosis and therapeutic approaches for each individual.

### Limitations

This study has some limitations inherent to its retrospective design. Despite standardised data collection, recall bias and variable clinical documentation may affect the accuracy of key variables like seizure history, precise recollection of milestone attainment, and age at treatment initiation. Missing data on treatment timing, duration, dosing, and milestone attainment further limit analysis precision. Additionally, neurodevelopmental outcomes were assessed primarily by clinicians, with only partial use of standardised neuropsychological tools, leading to potential inconsistencies. Even though randomised controlled trials would be most compelling, the rarity of the disease and the established efficacy of SCBs draws their feasibility into question.

Next, many individuals received a combination of different ASMs, making it challenging to isolate the effects of specific drugs on seizures. While we sought to address this by analysing the effect of each ASM according to its time of introduction, we did not account for precise dosages or therapeutic blood levels, nor an analysis of their concomitant medication. This lack of dosing and pharmacokinetic data makes it difficult to confirm optimal treatment exposure and may have resulted in both under- and overestimation of efficacy. Nevertheless, these limitations were present across all comparison groups, contributing to a degree of balance in the analysis.

The small sample size of the GOF subgroup limited our ability to evaluate the effects of ASMs—particularly since not all individuals experienced seizures. Larger studies in GOF cohorts will be necessary to clarify these issues. In addition, only individuals with functionally proven GOF variants were categorised in this subgroup. As not all missense variants were functionally tested, we cannot exclude the possibility of including an individual with a GOF variant in the LOF-DEE group, especially because individuals with GOF variants may present with non-epileptic myoclonic events during the neonatal period, which can be mistaken for seizures. All three individuals with post-neonatal seizure onset in the LOF-DEE group carried a proven dominant-negative LOF variant.

Despite these limitations, this study benefits from comprehensive, multinational data collection, and a large sample size relative to previous *KCNQ2* studies. The findings highlight clinically meaningful outcomes that can inform current practice and future research. Future, prospective, multicentre studies, ideally with standardised, and formal neurodevelopmental assessments are needed to confirm these findings and establish causality. Greater attention to therapeutic drug monitoring will be essential for optimising outcomes. In parallel, evaluating the impact of targeted therapies in well-characterised populations will be crucial for further improving prognosis in *KCNQ2*-related disorders.

### Conclusion and clinical implications

Our results strongly support the use of SCBs as first-line therapy in (LOF) *KCNQ2*-DEE and SeL(F)NE due to their high effectiveness. Moreover, SCBs appear most beneficial when initiated during the neonatal period, with earlier treatment linked to earlier seizure offset and better developmental outcomes. Given that treatment timing influences developmental trajectories, early genetic diagnosis is essential. Since unexplained neonatal-onset tonic or sequential seizures (i.e. after exclusion of the typical and common causes of neonatal seizures) are a hallmark of LOF *KCNQ2*-related disorders or other channelopathies shown to respond to SCB,^44,45^ physicians should not delay the initiation of SCB therapy when a genetic neonatal epilepsy is suspected, particularly if phenobarbital has proven ineffective. These data should inform future clinical guidelines, highlighting the need for genotype-specific precision therapy in neonatal epilepsy with a genetic aetiology.

## Data availability

All data supporting the findings of this study are available to those eligible upon request to the corresponding author.

## Acknowledgements

We thank the participating children and their parents, as well as the supporting parent organisations: European *KCNQ2* Association, *KCNQ2* e.V. (Germany), *KCNQ2* Cure Alliance, and *KCNQ2* France Développement. We also thank all the clinical research associates for their help and participation in the study, and in particular Chaimae Nadim for her valuable contribution in France. We thank Heiko Kienzler for support in data acquisition and curation.

## Supplementary material

Supplementary material is available at *Brain* online.

## Appendix

*KCNQ2* study group: Pasquale Striano, Francesca Madia, Rikke Steensbjerre Møller, Katrine M Johannesen, Ilona Krey, Oliver Maier, Astrid Bertsche, Pia Zacher, Dagmar Wieczorek, Margarete Koch-Hogrebe, Tobias Bartolomaeus, Lara Heuft, Marion Heruth, Janina Gburek-Augustat, Rebecca Gembicki, Irina Hüning, Malte Spielmann, Nienke Verbeek, Ernie Bongers, Christine Makowski, Berten Ceulemans, An-Sofie Schoonjans, Lieven Lagae, Mathias De Wachter, Katrien Jansen, Nina Barisic, Filip Roelens, Helene Verhelst, Ana Vilan, Johanneke Harteman, Liesbeth De Waele, Damien Lederer, Antonio Gil-Nagel, Paloma Parra-Díaz, Hilde Van Esch, Katrien Vanrykel, Jana Zarubova, Krystyna Szymanska, Jürgen Althaus, González Gutiérrez-Solana, Giulia Ventura, Pegoraro Veronica, Stephanie Colling, Britta Hanker, Celina Stülpnagel, Dorota Domańska-Pakieła, Angelo Russo, Joerg Klepper, Agnes Scheuring, Esther Mutz, Eulalia Turon-Viñas, Ivon Cuscó, Carlotta Stipa, Luca Bergonzini, Raffaella Minardi, Valentina Marchiani, Sophie Rus-Hall, Shannyn Genders, Bronwyn Grinton, Elena Pavlidis, Silvia Jorge, Anna Katharina Schönlaub, Susann Badmann, Andrea Berger, Matias Wagner, Elisabeth Cats, Michael Gößler, Nicholas M. Allen, Mary O’Regan, Preeyal Valla, Eoin P. Donnellan, Niamh McSweeney, Cian Duggan, Kathleen Gorman, Geoffroy Delplancq, Samer Wehbi, Tania Barragán**-**Arévalo, Gerhard Josef Kluger, Vitchayaporn Emarach Saengow, Céline Bovin, Fien Vandenberk, Eline Leermakers, Sylvie Lamoureux, Diana Rodriguez, Claude Cances, Maryline Carneiro, Camille Thomas, Marie-Laure Mathieu, Florence Robin-Ronaldo, Marie Christine Nougues, Helène Maurey, Sylvie Nguyen, Marie Vermelle, Adeline Trauffler, Emmanuel Cheuret, Benjamin Serrand, Julien Neveu, Christian Richelme, Rachel Froget, Agathe Roubertie, Thomas Smol, Sandrine Mary, Shannyn Genders, Nishtha Joshi

