## Supplementary Materials for "Early sodium channel blocker use improves seizures and neurodevelopment in *KCNQ2*-related disorders"

### Supplementary Table 1: Variant classification criteria for phenotype prediction in *KCNQ2*-related epilepsies created by the RIKEE *KCNQ2* expert panel

|  | ***KCNQ2-*related Self-limiting (familial) epilepsy**  **(SeLFNS/SeLFNIS/SeLFIS, +/- Myokymia)**  **OMIM** **121200:  Benign Familial Neonatal Seizures 1** | | | ***KCNQ2-*developmental and epileptic encephalopathy**  **(*KCNQ2*-DEE)**  **OMIM 613720: Developmental and epileptic encephalopathy 7** | | |
| --- | --- | --- | --- | --- | --- | --- |
|  | **Strong** | **Moderate** | **Supporting** | **Supporting** | **Moderate** | **Strong** |
| **Reported patient data** |  | Variant previously identified in a patient/pedigree with phenotype characteristic for self-limiting *KCNQ2* related epilepsy  **S*M1***  2 or more family members with self-limited neonatal epilepsy and normal development ***SM2*** |  | Variant identified in two unrelated clinical cases of DEE ***EP1*** | Variant identified in three unrelated clinical cases of DEE with incomplete clinical descriptions ***EM1*** | Variant identified in four or more unrelated clinical cases of DEE with incomplete clinical description or 3, if clinical descriptions are detailed and compatible with *KCNQ2*-DEE ***ES1*** |
| **Computational and Predictive Data** | Truncating variant expected to prevent expression of a subunit capable of tetramerization and self-limited phenotype ***SVS1*** (very strong)  Same variant previously identified in another pedigree with self-limited *KCNQ2* related epilepsy with 2 or more affected ***SS1***  Truncating variant expected to prevent expression of a subunit capable of tetramerization ***SS2***  Frameshift occurring beyond C-terminal D helix and self-limited phenotype ***SS3*** | Same amino acid change (different nucleotide change) as in an established pathogenic self-limited *KCNQ2* related epilepsy variant ***SM3*** | Novel missense change resulting in a conservative AA change outside one of 5 mutational hot spot domains for *KCNQ2*-DEE (S4-pore-C1-C4-C5) and no history of clinical encephalopathy/ developmental delay in index patient ***SP1*** | Novel missense change resulting in a non-conservative AA change at an AA residue where a different non-conservative AA change has been seen before in DEE or a conservative change has resulted in SeLFNE  ***EP2*** | Same amino acid change (different nucleotide change) as in an established pathogenic (DEE) variant ***EM2***  Missense or small in-frame deletion located in a transmembrane region outside one of 5 mutational hot spot domains for *KCNQ2*-DEE (S4-pore-C1, CaM B, link to SID, C-helix) and clinical encephalopathy/ developmental delay in index patient ***EM3*** | Missense or small in-frame deletion located within one of 5 mutational hot spot domains for *KCNQ2* -DEE (S4-pore-C1, CaM B, link to SID, C-helix) and clinical encephalopathy/ developmental delay in index patient ***ES2*** |
| **Functional Data** | Comprehensive electrophysiological studies show a modest (20-50%) loss of current  ***SS4*** |  | Limited electrophysiological study suggests a modest effect ***SP2*** |  | Comprehensive electrophysiological studies show a dominant-negative deleterious effect ***EM4***  Comprehensive electrophysiological studies show a gain-of-function effect ***EM5*** |  |
| **Family Data** | Large pedigree, 3 or more affected and good outcome in 3 generations, or 6 or more in total ***SVS2*** (very strong)  Moderate to large pedigree, 3 or more affected and good outcome ***SS5*** |  |  |  |  |  |
| **De novo occurrence** |  |  |  | De novo mutation and encephalopathic exam or delay in index patient, parents neurologically normal ***EP3*** | Confirmed mosaic parent with milder phenotype and infant with DEE ***EM6*** |  |
| **Clinical Data** |  | Index patient’s neurological exam and development is normal (in larger families, minority may be milder range impairments including recurrence of seizures) ***SM4*** |  | Index neonate’s neurological exam is encephalopathic and EEG is burst suppression; or, developmental delay in older index infants/children ***EP4*** |  |  |
|  | **SeLFNE**  Minimal   - 1 Very strong ***(SVS1-2)*** - 2 Strong **(SS1-5)** - 1 Strong ***(SS1-5)*** + 2 Moderate ***(SM1-4)*** - 1 Strong ***(SS1-5)*** + 1 Moderate ***(SM1-4)*** + 1 Supporting ***(SP1-2)*** | | | **DEE**  Minimal:   - 2 Strong ***(ES1-2)*** - 1 Strong ***(ES1-2)*** + 3 Moderate ***(EM1-6)*** - 1 Strong ***(ES1-2)*** + 2 Moderate ***(EM1-6)*** + 2 Supporting ***(EP1-4)*** - 1 Strong ***(ES1-2)*** + 1 Moderate ***(EM1-6)*** + 3 Supporting ***(EP1-4)*** | | |
|  | **Likely SeLFNE-related *KCNQ2* variant**  Minimal:   - 1 Strong ***(SS1-5)*** + 1 Moderate ***(SM1-4)*** - 1 Strong ***(SS1-5)*** + 2 Supporting ***(SP1-2)*** - 3 Moderate ***(SM1-4)*** - 2 Moderate + 2 Supporting ***(SP1-2)*** | | | **Likely DEE-related *KCNQ2* variant**  Minimal:   - 1 Strong ***(ES1-2)*** + 1 Moderate ***(EM1-6)*** - 1 Strong ***(ES1-2)*** + 2 Supporting ***(EP1-4)*** - 3 Moderate ***(EM1-6)*** - 2 Moderate ***(EM1-6)*** + 2 Supporting ***(EP1-4)*** | | |
|  | **Uncertain severity**  Any other combination | | | | | |

 AA = Amino acid; DEE = developmental and epileptic encephalopathy; EEG = electroencephalogram; SeLFIS = self-limited familial infantile seizures; SeLFNE  = self-limited familial neonatal epilepsy; SeLFNIS = self-limited familial neonatal and infantile seizures; SeLFNS = self-limited familial neonatal seizures

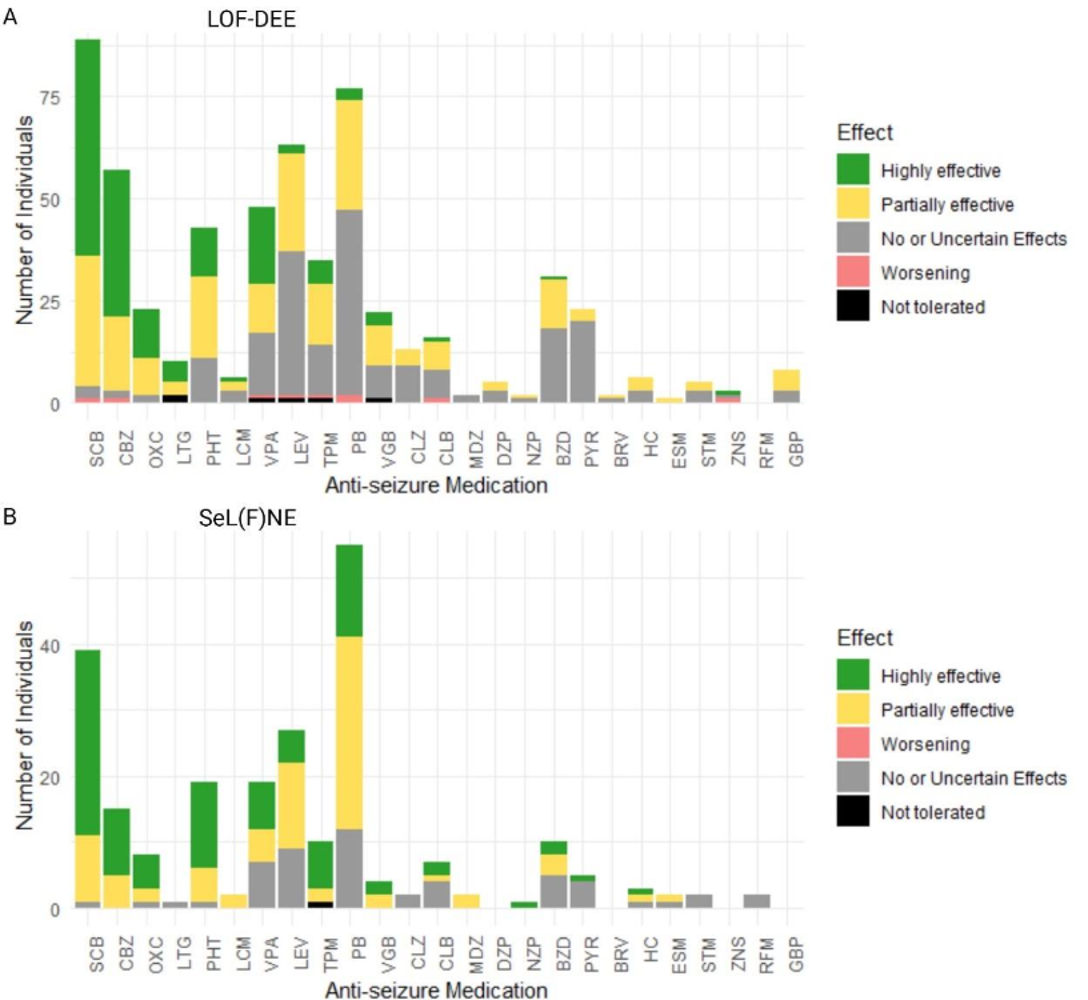

### **Supplementary Figure 1: Effects of all used antiseizure medications (ASMs) on seizures in LOF-DEE and SeL(F)NE cohorts.**

The figure shows reported treatment effects of ASMs among individuals with (likely) pathogenic *KCNQ2* variants predicted to lead to (A) loss-of-function developmental and epileptic encephalopathy (LOF-DEE) and (B) self-limited (familial) neonatal epilepsy (SeL(F)NE). Effects are categorised as highly effective (>90% seizure reduction), partially effective (reduction in seizure frequency between 50% and 90%), worsening (increase in seizure frequency), no or uncertain effects, and not tolerated. The stacked bar plots summarise the number of individuals experiencing each effect per ASM. For the category labeled 'SCB' (sodium channel blockers), the data represents a composite effect reflecting the best reported response among the individual SCB ASMs (CBZ, OXC, LTG, PHT, and LCM) administered. For example, if an individual experienced a partial effect with phenytoin (PHT) but a highly effective response with carbamazepine (CBZ), the 'SCB' category records the highest effect (i.e., highly effective) for that individual. This approach consolidates the overlapping mechanisms of these drugs in the analysis. Similarly, the 'BZD' (benzodiazepines) category summarises the combined effects of all benzodiazepines used by each individual, reflecting the overall BZD treatment effect regardless of specific drug. BRV = brivaracetam; BZD = benzodiazepines; CBZ = carbamazepine; CLB = clobazam; CLZ = clonazepam; DZP = diazepam; ESM = ethosuximide; FBM = felbamate; GBP = gabapentin; HC = hydrocortisone; LCM = lacosamide; LEV = levetiracetam; LOF-DEE = loss-of-function developmental and epileptic encephalopathy; LTG = lamotrigine; MDZ = midazolam; NZP = nitrazepam; OXC = oxcarbazepine; PB = phenobarbital; PHT = phenytoin; PER = perampanel; PYR = pyridoxin; RFM = rufinamide; SeL(F)NE = self-limited (familial) neonatal epilepsy; SCB = sodium channel blockers; STM = stiripentol; TPM = topiramate; VGB : vigabatrin; VPA = valproate; ZNS = zonisamide.

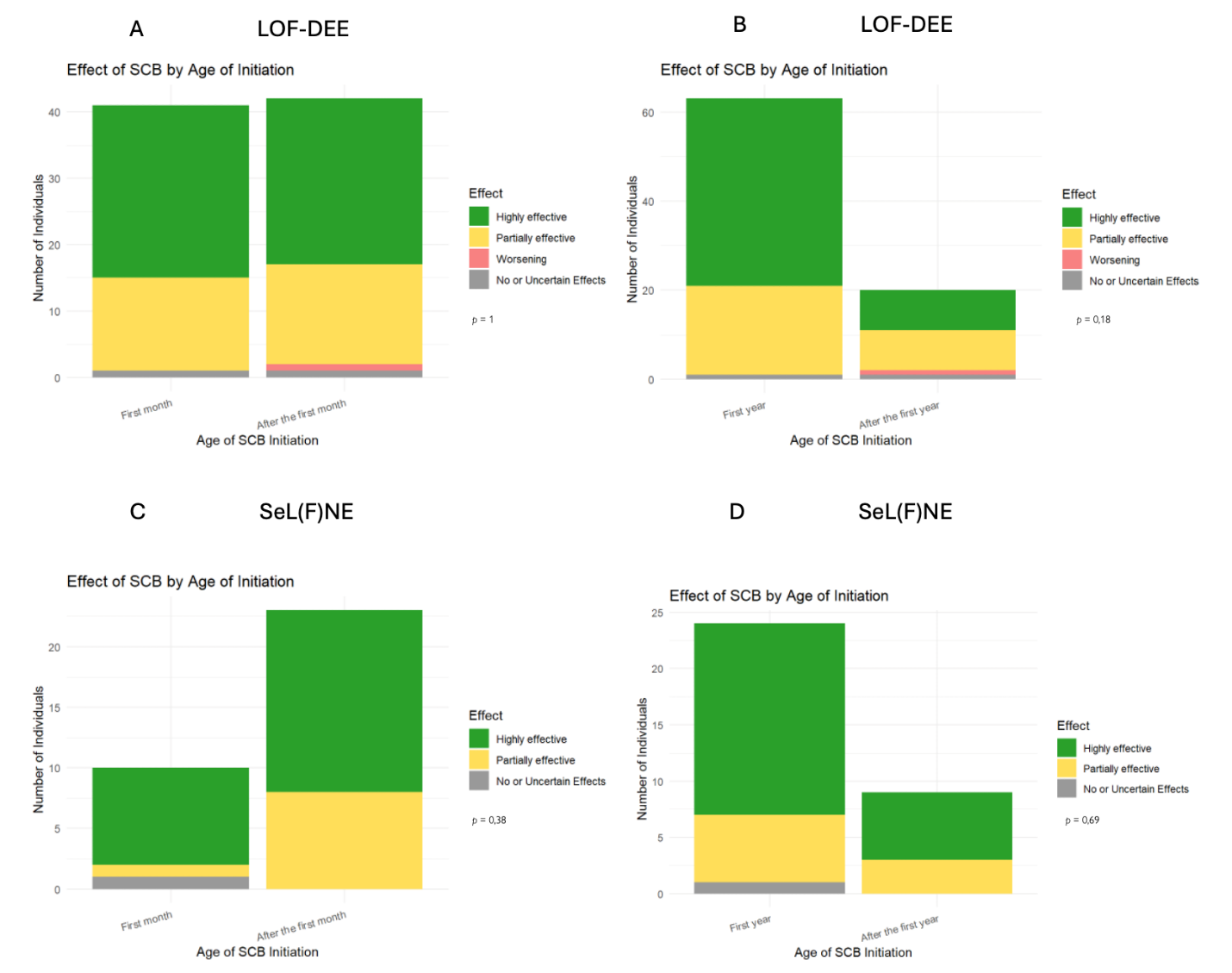

### **Supplementary Figure 2: Effects of sodium channel blocker (SCB) on seizures in LOF-DEE and SeL(F)NE cohorts**

The figure shows reported treatment effects of SCB with (likely) pathogenic KCNQ2 variants predicted to lead to (**A, B**) loss-of-function developmental and epileptic encephalopathy (LOF-DEE) and (**C, D**) self-limited (familial) neonatal epilepsy (SeL(F)NE). Different periods of SCB introduction were compared: before and after 1 month of age (**A, C**) and before and after 1 year of age (**B, D**). Effects are categorised as highly effective (>90% seizure reduction), partially effective (reduction in seizure frequency between 50% and 90%), worsening (increase in seizure frequency), no or uncertain effects, and not tolerated. The stacked bar plots summarise the number of individuals reporting the effects of SCBs. Reported treatment effects of SCBs were not significantly different when initiated before or after one month/year of life. (**A–D**) (Chi-square test, all *P* ≥ 0.05).
LOF-DEE = loss-of-function developmental and epileptic encephalopathy; SeL(F)NE = self-limited (familial) neonatal epilepsy; SCB = sodium channel blocker.

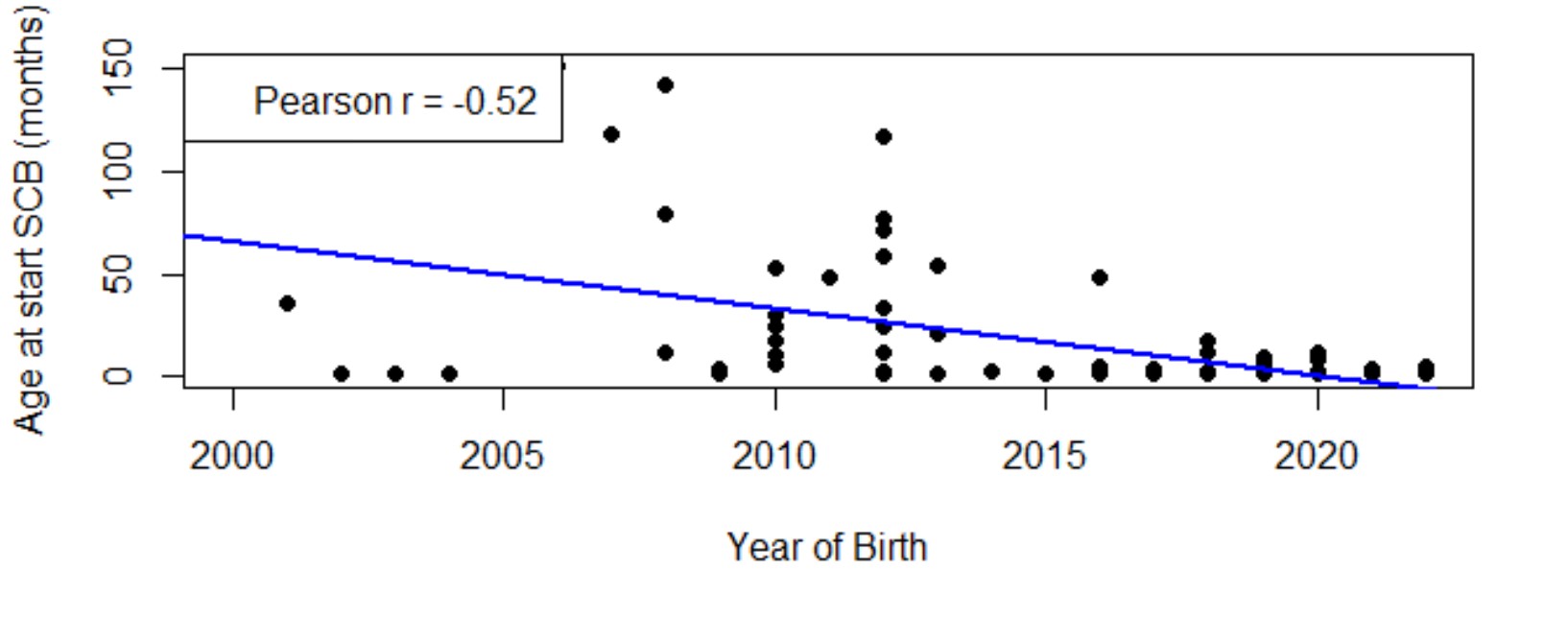

### **Supplementary Figure 3: Correlation between age at sodium channel blocker (SCB) initiation and year of birth in individuals with *KCNQ2* loss-of-function variants**

Scatter plots depicting the relationship between age at SCB initiation and year of birth, demonstrating a moderate negative correlation with a significant linear regression (Pearson r = -0.52, *P* < 0.0001). Blue regression lines indicate the fitted linear model. Pearson correlation coefficients (r) are included in each plot legend.

### Supplementary Table 2: Neurodevelopmental milestones and outcomes stratified by timing of sodium channel blocker initiation, using a 2-month of age cut-off, in individuals with *KCNQ2*-LOF DEE variants

|  | SCB initiation during first 2 months of life (n =43) | SCB initiation after second month of life (n =31) | Never taken SCBs (n =22) | P-value |
| --- | --- | --- | --- | --- |
| Median duration SCB use (months) (IQ1, IQ3) | 40 (16.5-59.8) | 53 (26-99) | NA |  |
| Achieved head control | 40/43 (93%) | 24/31 (77%) | 17/22 (77%) | 0.10***** |
| Head control   - < 4 months - > 4 months - Never | 7  10  3 | 1  10  7 | 0  5  5 | **0.03*** |
| Median age of head control (months) (IQ1, IQ3) | 4 (3–8) (n = 17) | 7 (5-11) (n =11) | 9 (6-12) (n =5) | 0.13**°** |
| Achieved independent sitting | 34/43 (79%) | 19/31 (61%) | 16/22 (73%) | 0.27***** |
| Sitting independently   - < 9 months - > 9 months - Never | 7  18  9 | 0  12  12 | 0  5  6 | **0.04*** |
| Median age of sitting independently (months) (IQ1, IQ3) | 12 (8 – 13) (n = 25) | 16 (10 – 19) (n = 12) | 13 (12 – 15) (n = 5) | 0.12° |
| Achieved independent walking | 30/43 (70%) | 18/31 (58%) | 14/22 (64%) | 0.58***** |
| Walking independently   - < 18 months - > 18 months - Never | 9  18  13 | 4  12  13 | 4  5  8 | 0.65* |
| Median age of walking independently (months) (IQ1, IQ3) | 22 (17.5 – 26.5) (n =27) | 29 (18.5 – 48.8) (n =16) | 20 (16 – 26) (n =9) | 0.12° |
| Achieved talking | 28/43 (65%) | 12/31 (39%) | 10/22 (45%) | 0.06* |
| First words   - < 16 months - > 16 months - Never | 8  10  15 | 2  8  19 | 3  4  12 | 0.35* |
| Median talking age (months) (IQ1, IQ3) | 17 (12.5 – 26) (n =19) | 28 (19 – 45) (n =10) | 18 (12 - 42) (n =7) | 0.18**°** |
| Neurodevelopmental outcome   - Normal - Mild/Moderate impairment - Severe/profound impairment | 7 (16%)  14 (33%)  22 (51%) | 1 (3%)  11 (35%)  19 (61%) | 1 (5%)  9 (41%)  12 (55%) | 0.34***** |

*Chi-square Test with Monte Carlo Simulation (B=10000); °Kruskal-Wallis rank sum test.

P-values are colour-coded to indicate statistical significance relative to SCB initiation timing groups (<1 month, >1 month, never): green denotes findings that maintain significance under this comparison, while red denotes findings that lose significance when analysed in this manner.

IQ1 = first interquartile; IQ3 = third interquartile; *KCNQ2*-LOF DEE = *KCNQ2*-related developmental and epileptic encephalopathy with loss-of-function; NA = not applicable; SCBs: sodium channel blockers

### Supplementary Table 3: Neurodevelopmental milestones and outcomes stratified by timing of sodium channel blocker initiation, using a 3-month of age cut-off, in individuals with *KCNQ2*-LOF DEE variants

|  | SCB initiation during first three months of life (n =46) | SCB initiation after third month of life (n =28) | Never taken SCBs (n =22) | P-value |
| --- | --- | --- | --- | --- |
| Median duration SCB use (months) (IQ1, IQ3) | 38.5 (16.2-59.4) | 63.5 (29.2-111) | NA |  |
| Achieved head control | 42/46 (91%) | 22/28 (79%) | 17/22 (77%) | 0.18***** |
| Head control   - < 4 months - > 4 months - Never | 7  10  4 | 1  10  6 | 0  5  5 | 0.06***** |
| Median age of head control (months) (IQ1, IQ3) | 4 (3–8) (n = 17) | 7 (5-11) (n =11) | 9 (6-12) (n =5) | 0.13**°** |
| Achieved independent sitting | 35/46 (76%) | 18/28 (64%) | 16/22 (73%) | 0.55***** |
| Sitting independently   - < 9 months - > 9 months - Never | 7  19  11 | 0  11  10 | 0  5  6 | 0.08***** |
| Median age of sitting independently (months) (IQ1, IQ3) | 11.5 (8.25-12.8) (n = 26) | 17 (11 – 20) (n = 11) | 13 (12 – 15) (n = 5) | 0.07° |
| Achieved independent walking | 31/46 (67%) | 17/28 (61%) | 14/22 (64%) | 0.85***** |
| Walking independently   - < 18 months - > 18 months - Never | 10  18  15 | 3  12  11 | 4  5  8 | 0.65***** |
| Median age of walking independently (months) (IQ1, IQ3) | 22 (16.8 – 26.2) (n =28) | 30 (21.5 – 49.5) (n =15) | 20 (16 – 26) (n =9) | **0.04°** |
| Achieved talking | 29/46 (63%) | 11/28 (39%) | 10/22 (45%) | 0.11* |
| First words   - < 16 months - > 16 months - Never | 9  10  17 | 1  8  17 | 3  4  12 | 0.23***** |
| Median talking age (months) (IQ1, IQ3) | 16.5 (12.8 – 25) (n =20) | 30 (22-48) (n =9) | 18 (12 - 42) (n =7) | 0.09**°** |
| Neurodevelopmental outcome   - Normal - Mild/Moderate impairment - Severe/profound impairment | 7 (15%)  15 (33%)  24 (52%) | 1 (4%)  10 (36%)  17 (61%) | 1 (5%)  9 (41%)  12 (55%) | 0.45***** |

*Chi-square Test with Monte Carlo Simulation (B=10000); °Kruskal-Wallis rank sum test.

P-values are colour-coded to indicate statistical significance relative to SCB initiation timing groups (<1 month, >1 month, never): red denotes findings that lose significance when analysed in this manner.

IQ1 = first interquartile; IQ3 = third interquartile; LOF-DEE = developmental and epileptic encephalopathy with loss-of-function variants; NA = not applicable; SCBs: sodium channel blockers

### Supplementary Table 4: Neurodevelopmental milestones and outcomes stratified by timing of sodium channel blocker initiation, using cut-offs of 1 month, 2–6 months, over 6 months or never, in individuals with *KCNQ2*-LOF DEE variants

|  | SCB initiation during first month of life (n =36) | SCB initiation between 2^nd^ and 6^th^ month of life (n =18) | SCB initiation after six months of life (n =20) | Never taken SCBs (n =22) | P-value |
| --- | --- | --- | --- | --- | --- |
| Median duration SCB use (months) (IQ1, IQ3) | 41 (16.8-67.5) | 32 (21.8-61.5) | 56.5 (26.5-121) | NA |  |
| Achieved head control | 35/36 (97%) | 15/18 (83%) | 14/20 (70%) | 17/22 (77%) | **0.03*** |
| Head control   - < 4 months - > 4 months - Never | 7  7  1 | 1  6  3 | 0  7  6 | 0  5  5 | **0.005*** |
| Median age of head control (months) (IQ1, IQ3) | 3.75 (3–5.75)  (n= 14) | 9 (7-20) (n =7) | 7 (5-9) (n =7) | 9 (6-12) (n =5) | **0.02°** |
| Achieved independent sitting | 32/36 (89%) | 9/18 (50%) | 12/20 (60%) | 16/22 (73%) | **0.01*** |
| Sitting independently   - < 9 months - > 9 months - Never | 7  16  4 | 0  7  9 | 0  7  8 | 0  5  6 | **0.005*** |
| Median age of sitting independently (months) (IQ1, IQ3) | 11 (7.5–12)  (n= 23) | 15 (11.5-23) (n =7) | 17 (11 – 18) (n = 7) | 13 (12 – 15) (n = 5) | 0.06° |
| Achieved independent walking | 29/36 (81%) | 8/18 (44%) | 11/20 (55%) | 14/22 (64%) | **0.04*** |
| Walking independently   - < 18 months - > 18 months - Never | 9  17  7 | 2  6  10 | 2  7  9 | 4  5  8 | 0.18* |
| Median age of walking independently (months) (IQ1, IQ3) | 22 (17.2–26.8)  (n=26) | 27 (18.5-38.2) (n =8) | 28 (24 – 51) (n =9) | 20 (16 – 26) (n =9) | 0.20° |
| Achieved talking | 25/36 (69%) | 7/18 (39%) | 8/20 (40%) | 10/22 (45%) | 0.13* |
| First words   - < 16 months - > 16 months - Never | 8  9  11 | 2  2  11 | 0  7  12 | 3  4  12 | 0.10* |
| Median talking age (months) (IQ1, IQ3) | 16.5 (12.2–24)  (n=18) | 28 (14-43.5) (n =4) | 30 (12 – 42) (n =7) | 18 (12 - 42) (n =7) | 0.14° |
| Neurodevelopmental outcome   - Normal - Mild/Moderate impairment - Severe/profound impairment | 7 (19%)  13 (36%)  16 (44%) | 1 (6%)  5 (28%)  12 (67%) | 0 (0%)  7 (35%)  13 (65%) | 1 (5%)  9 (41%)  12 (55%) | 0.20* |

**Chi-square Test with Monte Carlo Simulation (B=10000); °Kruskal-Wallis rank sum test.

P-values are colour-coded to indicate statistical significance relative to SCB initiation timing groups (<1 month, >1 month, never): green denotes findings that maintain significance under this comparison, while red denotes findings that lose significance when analysed in this manner.

IQ1 = first interquartile; IQ3 = third interquartile; *KCNQ2*-LOF DEE = *KCNQ2*-related developmental and epileptic encephalopathy with loss-of-function; NA = not applicable; SCBs: sodium channel blockers

### Supplementary Table 5: Neurodevelopmental milestones and outcomes stratified by timing of seizure offset, using a 2-month age threshold, in individuals with *KCNQ2*-LOF DEE variants

|  | Seizure offset during the first two months of life (n =34) | Seizure offset after the second month of life (n =64) | Never experienced a period of seizure offset (n =10) | P-value |
| --- | --- | --- | --- | --- |
| Achieved head control | 33/34 (97%) | 52/64 (81%) | 2/10 (20%) | **<0.0001*** |
| Head control   - < 4 months - > 4 months - Never | 6  7  1 | 3  18  12 | 0  1  8 | **0.0006*** |
| Median age of head control (months) (IQ1, IQ3) | 4 (3 – 6) (n = 13) | 8 (5 – 12) (n =21) | 7 (7-7) (n =1) | 0.14° |
| Achieved independent sitting | 28/34 (82%) | 46/64 (72%) | 1/10 (10%) | **<0.0001*** |
| Sitting independently   - < 9 months - > 9 months - Never | 5  11  6 | 2  26  18 | 0  0  9 | **0.001** |
| Median age of sitting independently (months) (IQ1, IQ3) | 12 (7 – 14) (n = 16) | 12.5 (10.8-19) (n = 28) | *NA* | 0.08**°** |
| Achieved independent walking | 24/34 (71%) | 43/64 (67%) | 1/10 (10%) | **0.001*** |
| Walking independently   - < 18 months - > 18 months - Never | 7  14  10 | 11  24  22 | 0  1  9 | **0.02** |
| Median age of walking independently (months) (IQ1, IQ3) | 21 (17 – 28) (n =21) | 24 (17.5 – 35.5) (n =35) | 24 (24-24) (n =1) | 0.62° |
| Achieved talking | 22/34 (65%) | 32/64 (50%) | 1/10 (10%) | **0.01*** |
| First words   - < 16 months - > 16 months - Never | 8  7  12 | 6  16  32 | 0  1  9 | **0.048** |
| Median talking age (months) (IQ1, IQ3) | 15 (12 – 32) (n =15) | 18 (15.5 – 39) (n =23) | 22 (22-22) (n =1) | 0.42° |
| Neurodevelopmental outcome   - Normal - Mild/Moderate impairment - Severe/profound impairment | 6 (18%)  11 (32%)  17 (50%) | 3 (5%)  26 (41%)  35 (55%) | 0 (0%)  1 (10%)  9 (90%) | **0.04*** |

*Chi-square Test with Monte Carlo Simulation (B=10000); °Kruskal-Wallis rank sum test.

P-values are colour-coded to indicate statistical significance relative to seizure offset timing groups (<1 month, >1 month, never): green denotes findings that maintain significance under this comparison, while red denotes findings that lose significance when analysed in this manner.

IQ1 = first interquartile; IQ3 = third interquartile; *KCNQ2*-LOF DEE = *KCNQ2*-related developmental and epileptic encephalopathy with loss-of-function

### Supplementary Table 6: Neurodevelopmental milestones and outcomes stratified by timing of seizure offset, using a 3-month age threshold, in individuals with *KCNQ2*-LOF DEE variants

|  | Seizure offset during the first three months of life (n =40) | Seizure offset after the third month of life (n =58) | Never experienced a period of seizure offset (n =10) | P-value |
| --- | --- | --- | --- | --- |
| Achieved head control | 39/40 (98%) | 46/58 (79%) | 2/10 (20%) | **<0.0001*** |
| Head control   - < 4 months - > 4 months - Never | 6  10  1 | 3  15  12 | 0  1  8 | **0.0006*** |
| Median age of head control (months) (IQ1, IQ3) | 4.5 (3 – 8.25) (n = 16) | 7.5 (5 – 14.2) (n =18) | 7 (7-7) (n =1) | 0.26° |
| Achieved independent sitting | 34/40 (85%) | 40/58 (69%) | 1/10 (10%) | **<0.0001*** |
| Sitting independently   - < 9 months - > 9 months - Never | 6  14  6 | 1  23  18 | 0  0  9 | **0.0002*** |
| Median age of sitting independently (months) (IQ1, IQ3) | 12 (7.75 – 17) (n = 20) | 12.5 (11-19) (n = 24) | *NA* | 0.13**°** |
| Achieved independent walking | 29/40 (73%) | 38/58 (66%) | 1/10 (10%) | **0.001*** |
| Walking independently   - < 18 months - > 18 months - Never | 9  16  11 | 9  22  20 | 0  1  9 | **0.02*** |
| Median age of walking independently (months) (IQ1, IQ3) | 20 (17 – 24) (n =25) | 26 (17.5-36) (n =31) | 24 (24-24) (n =1) | 0.27° |
| Achieved talking | 27/40 (68%) | 27/58 (47%) | 1/10 (10%) | **0.002*** |
| First words   - < 16 months - > 16 months - Never | 9  9  13 | 5  14  31 | 0  1  9 | **0.03*** |
| Median talking age (months) (IQ1, IQ3) | 19.5 (12 – 27.5) (n =18) | 18 (15.8 – 43.5) (n =20) | 22 (22-22) (n =1) | 0.39° |
| Neurodevelopmental outcome   - Normal - Mild/Moderate impairment - Severe/profound impairment | 7 (18%)  15 (38%)  18 (45%) | 2 (3%)  22 (38%)  34 (59%) | 0 (0%)  1 (10%)  9 (90%) | **0.02*** |

*Chi-square Test with Monte Carlo Simulation (B=10000); °Kruskal-Wallis rank sum test.

P-values are colour-coded to indicate statistical significance relative to seizure offset timing groups (<1 month, >1 month, never): green denotes findings that maintain significance under this comparison, while red denotes findings that lose significance when analysed in this manner.

IQ1 = first interquartile; IQ3 = third interquartile; *KCNQ2*-LOF DEE = *KCNQ2*-related developmental and epileptic encephalopathy with loss-of-function; NA = not applicable

### Supplementary Table 7: Neurodevelopmental milestones and outcomes stratified by timing of seizure offset, using a 4-month age cut-off, in individuals with *KCNQ2*-LOF DEE variants

|  | Seizure offset during the first four months of life (n =48) | Seizure offset after the 4th month of life (n =50) | Never experienced a period of seizure offset (n =10) | P-value |
| --- | --- | --- | --- | --- |
| Achieved head control | 45/48 (94%) | 40/50 (80%) | 2/10 (20%) | **<0.0001*** |
| Head control   - < 4 months - > 4 months - Never | 7  11  3 | 2  14  10 | 0  1  8 | **0.0007** |
| Median age of head control (months) (IQ1, IQ3) | 4.5 (3 – 7.5) (n = 18) | 8.5 (5 – 15) (n =16) | 7 (7-7) (n =1) | 0.10° |
| Achieved independent sitting | 40/48 (83%) | 34/50 (68%) | 1/10 (10%) | **<0.0001*** |
| Sitting independently   - < 9 months - > 9 months - Never | 6  18  8 | 1  19  16 | 0  0  9 | **0.0005** |
| Median age of sitting independently (months) (IQ1, IQ3) | 12 (8.75 – 17.2) (n = 24) | 12 (11-19) (n = 20) | *NA* | 0.34**°** |
| Achieved independent walking | 35/48 (73%) | 32/50 (64%) | 1/10 (10%) | **0.0007*** |
| Walking independently   - < 18 months - > 18 months - Never | 10  20  13 | 8  18  18 | 0  1  9 | **0.02** |
| Median age of walking independently (months) (IQ1, IQ3) | 21.5 (17.2 – 27.8) (n =30) | 26 (17.2-35.8) (n =26) | 24 (24-24) (n =1) | 0.64° |
| Achieved talking | 32/48 (67%) | 22/50 (44%) | 1/10 (10%) | **0.002*** |
| First words   - < 16 months - > 16 months - Never | 9  12  16 | 5  11  28 | 0  1  9 | 0.06* |
| Median talking age (months) (IQ1, IQ3) | 24 (12 – 36) (n =21) | 18 (15 – 48) (n =17) | 22 (22-22) (n =1) | 0.71° |
| Neurodevelopmental outcome   - Normal - Mild/Moderate impairment - Severe/profound impairment | 7 (15%)  20 (42%)  21 (44%) | 2 (4%)  17 (34%)  31 (62%) | 0 (0%)  1 (10%)  9 (90%) | **0.03*** |

*Chi-square Test with Monte Carlo Simulation (B=10000); °Kruskal-Wallis rank sum test.

P-values are colour-coded to indicate statistical significance relative to seizure offset timing groups (<1 month, >1 month, never): green denotes findings that maintain significance under this comparison, while red denotes findings that lose significance when analysed in this manner.

IQ1 = first interquartile; IQ3 = third interquartile; *KCNQ2*-LOF DEE = *KCNQ2*-related developmental and epileptic encephalopathy with loss-of-function; NA = not applicable

### Supplementary Table 8: Neurodevelopmental milestones and outcomes stratified by timing of seizure offset, using a 6-month age cut-off, in individuals with *KCNQ2*-LOF DEE variants

|  | Seizure offset during the first six months of life (n =57) | Seizure offset after the 6th month of life (n =41) | Never experienced a period of seizure offset (n =10) | P-value |
| --- | --- | --- | --- | --- |
| Achieved head control | 54/57 (95%) | 31/41 (76%) | 2/10 (20%) | **<0.0001*** |
| Head control   - < 4 months - > 4 months - Never | 8  14  3 | 1  11  10 | 0  1  8 | **0.0005** |
| Median age of head control (months) (IQ1, IQ3) | 5 (3 – 8.75) (n = 22) | 8.5 (5.75 – 12.8) (n =12) | 7 (7-7) (n =1) | 0.20° |
| Achieved independent sitting | 48/57 (84%) | 26/41 (63%) | 1/10 (10%) | **<0.0001*** |
| Sitting independently   - < 9 months - > 9 months - Never | 6  24  9 | 1  13  15 | 0  0  9 | **0.0007** |
| Median age of sitting independently (months) (IQ1, IQ3) | 12 (9.25 – 17.8) (n = 30) | 12 (11-21) (n = 14) | *NA* | 0.49**°** |
| Achieved independent walking | 43/57 (75%) | 24/41 (59%) | 1/10 (10%) | **0.0007*** |
| Walking independently   - < 18 months - > 18 months - Never | 11  27  14 | 7  11  17 | 0  1  9 | **0.004** |
| Median age of walking independently (months) (IQ1, IQ3) | 23 (18 – 33) (n =38) | 25 (17-35.2) (n =18) | 24 (24-24) (n =1) | 0.99° |
| Achieved talking | 37/57 (65%) | 17/41 (41%) | 1/10 (10%) | **0.002*** |
| First words   - < 16 months - > 16 months - Never | 10  15  20 | 4  8  24 | 0  1  9 | 0.06 |
| Median talking age (months) (IQ1, IQ3) | 18 (13 – 36) (n =25) | 18 (15 – 60) (n =13) | 22 (22-22) (n =1) | 0.68° |
| Neurodevelopmental outcome   - Normal - Mild/Moderate impairment - Severe/profound impairment | 8 (14%)  24 (42%)  25 (44%) | 1 (2%)  13 (32%)  27 (66%) | 0 (0%)  1 (10%)  9 (90%) | **0.02*** |

*Chi-square Test with Monte Carlo Simulation (B=10000); °Kruskal-Wallis rank sum test.

P-values are colour-coded to indicate statistical significance relative to seizure offset timing groups (<1 month, >1 month, never): green denotes findings that maintain significance under this comparison, while red denotes findings that lose significance when analysed in this manner.

IQ1 = first interquartile; IQ3 = third interquartile; *KCNQ2*-LOF DEE = *KCNQ2*-related developmental and epileptic encephalopathy with loss-of-function; NA = not applicable

### Supplementary Table 9: Neurodevelopmental milestones and outcomes stratified by timing of seizure offset, using an 8-month age cut-off, in individuals with *KCNQ2*-LOF DEE variants

|  | Seizure offset during the first eight months of life (n =60) | Seizure offset after the 8th month of life (n =38) | Never experienced a period of seizure offset (n =10) | P-value |
| --- | --- | --- | --- | --- |
| Achieved head control | 57/60 (95%) | 28/38 (74%) | 2/10 (20%) | **<0.0001*** |
| Head control   - < 4 months - > 4 months - Never | 9  15  3 | 0  10  10 | 0  1  8 | **<0.0001*** |
| Median age of head control (months) (IQ1, IQ3) | 4.5 (3 – 8.25) (n = 24) | 10.5 (7.25 – 14.2) (n =10) | 7 (7-7) (n =1) | **0.02°** |
| Achieved independent sitting | 50/60 (83%) | 24/38 (63%) | 1/10 (10%) | **<0.0001*** |
| Sitting independently   - < 9 months - > 9 months - Never | 6  26  10 | 1  11  14 | 0  0  9 | **0.0009** |
| Median age of sitting independently (months) (IQ1, IQ3) | 12 (9.75 – 17.2) (n = 32) | 12 (11-23.2) (n = 12) | *NA* | **0.36°** |
| Achieved independent walking | 45/60 (75%) | 22/38 (58%) | 1/10 (10%) | **0.0005*** |
| Walking independently   - < 18 months - > 18 months - Never | 11  28  15 | 7  10  16 | 0  1  9 | **0.003** |
| Median age of walking independently (months) (IQ1, IQ3) | 24 (18 – 32) (n =39) | 24 (17-36) (n =17) | 24 (24-24) (n =1) | 0.99° |
| Achieved talking | 39/60 (65%) | 15/38 (39%) | 1/10 (10%) | **0.0009*** |
| First words   - < 16 months - > 16 months - Never | 10  15  21 | 4  8  23 | 0  1  9 | **0.08** |
| Median talking age (months) (IQ1, IQ3) | 18 (13 – 36) (n =25) | 18 (15 – 60) (n =13) | 22 (22-22) (n =1) | 0.68° |
| Neurodevelopmental outcome   - Normal - Mild/Moderate impairment - Severe/profound impairment | 8 (13%)  26 (43%)  26 (43%) | 1 (3%)  11 (29%)  26 (68%) | 0 (0%)  1 (10%)  9 (90%) | **0.02*** |

*Chi-square Test with Monte Carlo Simulation (B=10000); °Kruskal-Wallis rank sum test.

P-values are colour-coded to indicate statistical significance relative to seizure offset timing groups (<1 month, >1 month, never): green denotes findings that maintain significance under this comparison, red denotes findings that lose significance when analysed in this manner.

IQ1 = first interquartile; IQ3 = third interquartile; *KCNQ2*-LOF DEE = *KCNQ2*-related developmental and epileptic encephalopathy with loss-of-function; NA = not applicable

### Supplementary Table 10: Neurodevelopmental milestones and outcomes stratified by timing of seizure offset, using a 12-month age cut-off, in individuals with *KCNQ2*-LOF DEE variants

|  | Seizure offset during the first year of life (n =71) | Seizure offset after the first year of life (n =27) | Never experienced a period of seizure offset (n =10) | P-value |
| --- | --- | --- | --- | --- |
| Achieved head control | 67/71 (94%) | 18/27 (67%) | 2/10 (20%) | **<0.0001*** |
| Head control   - < 4 months - > 4 months - Never | 9  21  4 | 0  4  9 | 0  1  8 | **0.0002*** |
| Median age of head control (months) (IQ1, IQ3) | 5 (3.12 – 9.75) (n = 30) | 6.5 (5.75 – 8.25) (n =4) | 7 (7-7) (n =1) | 0.83° |
| Achieved independent sitting | 59/71 (83%) | 15/27 (56%) | 1/10 (10%) | **<0.0001*** |
| Sitting independently   - < 9 months - > 9 months - Never | 6  31  12 | 1  6  12 | 0  0  9 | **0.0003** |
| Median age of sitting independently (months) (IQ1, IQ3) | 12 (10 – 18) (n = 37) | 11 (10.5-12) (n = 7) | *NA* | 0.38**°** |
| Achieved independent walking | 53/71 (75%) | 14/27 (52%) | 1/10 (10%) | **0.0002*** |
| Walking independently   - < 18 months - > 18 months - Never | 14  33  18 | 4  5  13 | 0  1  9 | **0.0011** |
| Median age of walking independently (months) (IQ1, IQ3) | 24 (17.5 – 35.5) (n =47) | 19 (16-26) (n =9) | 24 (24-24) (n =1) | 0.69° |
| Achieved talking | 45/71 (63%) | 9/27 (33%) | 1/10 (10%) | **0.0008*** |
| First words   - < 16 months - > 16 months - Never | 12  18  26 | 2  5  18 | 0  1  9 | **0.04** |
| Median talking age (months) (IQ1, IQ3) | 21 (13.2 – 34.5) (n =30) | 18 (15.5 – 102) (n =8) | 22 (22-22) (n =1) | 0.80° |
| Neurodevelopmental outcome   - Normal - Mild/Moderate impairment - Severe/profound impairment | 8 (11%)  32 (45%)  31 (44%) | 1 (4%)  5 (19%)  21 (78%) | 0 (0%)  1 (10%)  9 (90%) | **0.009*** |

*Chi-square Test with Monte Carlo Simulation (B=10000); °Kruskal-Wallis rank sum test.

P-values are colour-coded to indicate statistical significance relative to seizure offset timing groups (<1 month, >1 month, never): green denotes findings that maintain significance under this comparison, while red denotes findings that lose significance when analysed in this manner.

IQ1 = first interquartile; IQ3 = third interquartile; *KCNQ2*-LOF DEE = *KCNQ2*-related developmental and epileptic encephalopathy with loss-of-function; NA = not applicable

### Supplementary Table 11: Neurodevelopmental milestones and outcomes stratified by timing of seizure offset, using age cut-offs of ≤3 months, >3–12 months, and >12 months or never, in individuals with *KCNQ2*-LOF DEE variants

|  | Seizure offset during the first three months of life (n =40) | Seizure offset after the third month and up to the twelfth month of life (n =31) | Seizure offset after 1 year or never (n =37) | P-value |
| --- | --- | --- | --- | --- |
| Achieved head control | 39/40 (98%) | 28/31 (90%) | 20/37 (54%) | **<0.0001*** |
| Head control   - < 4 months - > 4 months - Never | 6  10  1 | 3  11  3 | 0  5  17 | **<0.0001*** |
| Median age of head control (months) (IQ1, IQ3) | 4.5 (3 – 8.25) (n = 16) | 8.5 (4.25 – 15) (n =14) | 7 (6-7) (n =5) | 0.26° |
| Achieved independent sitting | 34/40 (85%) | 25/31 (81%) | 16/37 (43%) | **<0.0001*** |
| Sitting independently   - < 9 months - > 9 months - Never | 6  14  6 | 0  17  6 | 1  6  21 | **<0.0001*** |
| Median age of sitting independently (months) (IQ1, IQ3) | 12 (7.75 – 17) (n = 20) | 15 (12-22) (n = 17) | 11 (10.5-12) (n =7) | 0.09**°** |
| Achieved independent walking | 29/40 (73%) | 24/31 (77%) | 15/37 (41%) | **0.003*** |
| Walking independently   - < 18 months - > 18 months - Never | 9  16  11 | 5  17  7 | 4  6  22 | **0.003** |
| Median age of walking independently (months) (IQ1, IQ3) | 20 (17 – 24) (n =25) | 28.5 (19.8-37.5) (n =22) | 21.5 (16.5-25.5) (n =10) | 0.08° |
| Achieved talking | 27/40 (68%) | 18/31 (58%) | 10/37 (27%) | **0.001*** |
| First words   - < 16 months - > 16 months - Never | 9  9  13 | 3  9  13 | 2  6  27 | **0.016** |
| Median talking age (months) (IQ1, IQ3) | 19.5 (12 – 27.5) (n =18) | 23.5 (16.5 – 37.5) (n =12) | 18 (16-96) (n =9) | 0.39° |
| Neurodevelopmental outcome   - Normal - Mild/Moderate impairment - Severe/profound impairment | 7 (18%)  15 (38%)  18 (45%) | 2 (6%)  17 (55%)  13 (42%) | 1 (3%)  6 (16%)  30 (81%) | **0.0009*** |

*Chi-square Test with Monte Carlo Simulation (B=10000); °Kruskal-Wallis rank sum test.

P-values are colour-coded to indicate statistical significance relative to seizure offset timing groups (<1 month, >1 month, never): green denotes findings that maintain significance under this comparison, while red denotes findings that lose significance when analysed in this manner.

IQ1 = first interquartile; IQ3 = third interquartile; *KCNQ2*-LOF DEE = *KCNQ2*-related developmental and epileptic encephalopathy with loss-of-function; NA = not applicable

### Supplementary Table 12: Neurodevelopmental milestones and outcomes stratified by timing of seizure offset, using age cut-offs of ≤2 months, >2–12 months, and >12 months or never, in individuals with *KCNQ2*-LOF DEE variants

|  | Seizure offset during the first two months of life (n =34) | Seizure offset after the second month and up to the twelfth month of life (n =37) | Seizure offset after 1 year or never (n =37) | P-value |
| --- | --- | --- | --- | --- |
| Achieved head control | 33/34 (97%) | 34/37 (92%) | 20/37 (54%) | **<0.0001*** |
| Head control   - < 4 months - > 4 months - Never | 6  7  1 | 3  14  3 | 0  5  17 | **<0.0001*** |
| Median age of head control (months) (IQ1, IQ3) | 4 (3 – 6) (n = 13) | 8 (5 – 15) (n =17) | 7 (6-7) (n =5) | 0.14° |
| Achieved independent sitting | 28/34 (82%) | 31/37 (84%) | 16/37 (43%) | **0.0002*** |
| Sitting independently   - < 9 months - > 9 months - Never | 5  11  6 | 1  20  6 | 1  6  21 | **<0.0001*** |
| Median age of sitting independently (months) (IQ1, IQ3) | 12 (7 – 14) (n = 16) | 15 (11-22) (n = 21) | 11 (10.5-12) (n =7) | 0.07**°** |
| Achieved independent walking | 24/34 (71%) | 29/37 (78%) | 15/37 (41%) | **0.003*** |
| Walking independently   - < 18 months - > 18 months - Never | 7  14  10 | 7  19  8 | 4  6  22 | **0.002** |
| Median age of walking independently (months) (IQ1, IQ3) | 21 (17 – 28) (n =21) | 26.5 (18.2-36) (n =26) | 21.5 (16.5-25.5) (n =10) | 0.31° |
| Achieved talking | 22/34 (65%) | 23/37 (62%) | 10/37 (27%) | **0.002*** |
| First words   - < 16 months - > 16 months - Never | 8  7  12 | 4  11  14 | 2  6  27 | **0.015** |
| Median talking age (months) (IQ1, IQ3) | 15 (12 – 32) (n =15) | 24 (16 – 33) (n =15) | 18 (16-96) (n =9) | 0.42° |
| Neurodevelopmental outcome   - Normal - Mild/Moderate impairment - Severe/profound impairment | 6 (18%)  11 (32%)  17 (50%) | 2 (5%)  21 (57%)  14 (38%) | 1 (3%)  6 (16%)  30 (81%) | **0.0006*** |

*Chi-square Test with Monte Carlo Simulation (B=10000); °Kruskal-Wallis rank sum test.

P-values are colour-coded to indicate statistical significance relative to seizure offset timing groups (<1 month, >1 month, never): green denotes findings that maintain significance under this comparison, while red denotes findings that lose significance when analysed in this manner.

IQ1 = first interquartile; IQ3 = third interquartile; *KCNQ2*-LOF DEE = *KCNQ2*-related developmental and epileptic encephalopathy with loss-of-function; NA = not applicable

### Supplementary Table 13: Neurodevelopmental milestones and outcomes stratified by timing of seizure offset, using age thresholds of ≤1 month, >1–12 months, and >12 months or never, in individuals with *KCNQ2*-LOF DEE variants

|  | Seizure offset during the first month of life (n =22) | Seizure offset after the first month and up to the twelfth month of life (n =49) | Seizure offset after 1 year or never (n =37) | P-value |
| --- | --- | --- | --- | --- |
| Achieved head control | 22/22 (100%) | 45/49 (92%) | 20/37 (54%) | **<0.0001*** |
| Head control   - < 4 months - > 4 months - Never | 6  3  0 | 3  18  4 | 0  5  17 | **<0.0001*** |
| Median age of head control (months) (IQ1, IQ3) | 3 (3 – 4) (n= 9) | 8 (5 – 15) (n =21) | 7 (6-7) (n =5) | **0.02°** |
| Achieved independent sitting | 20/22 (91%) | 39/49 (80%) | 16/37 (43%) | **<0.0001*** |
| Sitting independently   - < 9 months - > 9 months - Never | 5  6  2 | 1  25  10 | 1  6  21 | **<0.0001*** |
| Median age of sitting independently (months) (IQ1, IQ3) | 9 (7 – 12) (n= 11) | 16 (12-21) (n = 26) | 11 (10.5-12) (n =7) | **0.007°** |
| Achieved independent walking | 19/22 (86%) | 34/49 (69%) | 15/37 (41%) | **0.0007*** |
| Walking independently   - < 18 months - > 18 months - Never | 6  10  3 | 8  23  15 | 4  6  22 | **0.001** |
| Median age of walking independently (months) (IQ1, IQ3) | 20.5 (15.8 – 24) (n=16) | 27 (18.5-40) (n =31) | 21.5 (16.5-25.5) (n =10) | 0.05° |
| Achieved talking | 17/22 (77%) | 28/49 (57%) | 10/37 (27%) | **0.0004*** |
| First words   - < 16 months - > 16 months - Never | 6  5  5 | 6  13  21 | 2  6  27 | **0.009** |
| Median talking age (months) (IQ1, IQ3) | 13 (12 – 27) (n=11) | 24 (15 – 39) (n =19) | 18 (16-96) (n =9) | 0.27° |
| Neurodevelopmental outcome   - Normal - Mild/Moderate impairment - Severe/profound impairment | 6 (27%)  8 (36%)  8 (36%) | 2 (4%)  24 (49%)  23 (47%) | 1 (3%)  6 (16%)  30 (81%) | **<0.0001*** |

*Chi-square Test with Monte Carlo Simulation (B=10000); °Kruskal-Wallis rank sum test.

P-values are colour-coded to indicate statistical significance relative to seizure offset timing groups (<1 month, >1 month, never): green denotes findings that maintain significance under this comparison, while red denotes findings that lose significance when analysed in this manner.

IQ1 = first interquartile; IQ3 = third interquartile; *KCNQ2*-LOF DEE = *KCNQ2*-related developmental and epileptic encephalopathy with loss-of-function; NA = not applicable

### Supplementary Table 14: Individual-level data in a subset of individuals carrying recurrent variants

| *KCNQ2* Variant | Subject | Early SCBs use | Early seizure offset | Neurodevelopmental outcome |
| --- | --- | --- | --- | --- |
| p.(Arg560Trp) | 1 | No | No | Mild to moderate |
| p.(Arg560Trp) | 2 | No | NA | Mild to moderate |
| p.(Arg560Trp) | 3 | Yes | No | Severe to profound |
| p.(Arg560Trp) | 4 | Yes | No | Severe to profound |
| p.(Arg560Trp) | 5 | Yes | No | Severe to profound |
| p.(Arg560Trp) | 6 | Yes | No | Severe to profound |
| p.(Arg560Trp) | 7 | No | No | Severe to profound |
| p.(Arg560Trp) | 8 | NA | No | Severe to profound |
| p.(Arg560Trp) | 9 | NA | No | Severe to profound |
| p.(Arg213Trp) | 1 | Yes | Yes | Normal |
| p.(Arg213Trp) | 2 | Yes | Yes | Normal |
| p.(Arg213Trp) | 3 | No | No | Mild to moderate |
| p.(Arg213Trp) | 4 | Yes | Yes | Severe to profound |
| p.(Arg213Trp) | 5 | Yes | No | Severe to profound |
| p.(Arg213Trp) | 6 | No | No | Severe to profound |
| p.(Arg213Trp) | 7 | NA | No | Severe to profound |
| p.(Arg213Trp) | 8 | NA | No | Severe to profound |
| p.(Ala196Val) | 1 | Yes | Yes | Normal |
| p.(Ala196Val) | 2 | Yes | No | Normal |
| p.(Ala196Val) | 3 | No | No | Normal |
| p.(Ala196Val) | 4 | NA | No | Mild to moderate |
| p.(Ala196Val) | 5 | No | No | Mild to moderate |
| p.(Ala265Thr) | 1 | No | No | Severe to profound |
| p.(Ala265Thr) | 2 | No | No | Severe to profound |
| p.(Ala265Thr) | 3 | No | No | Severe to profound |
| p.(Ala265Thr) | 4 | NA | No | Severe to profound |
| p.(Ala265Val) | 1 | Yes | Yes | Normal |
| p.(Ala265Val) | 2 | Yes | No | Mild to moderate |
| p.(Ala265Val) | 3 | No | No | Mild to moderate |
| p.(Ala265Val) | 4 | No | No | Severe to profound |
| p.(Arg207Trp) | 1 | Yes | Yes | Mild to moderate |
| p.(Arg207Trp) | 2 | No | No | Mild to moderate |
| p.(Arg207Trp) | 3 | No | No | Mild to moderate |
| p.(Arg207Trp) | 4 | No | No | Mild to moderate |
| p.(Ala306Val) | 1 | Yes | No | Mild to moderate |
| p.(Ala306Val) | 2 | No | No | Mild to moderate |
| p.(Ala306Val) | 3 | NA | No | Mild to moderate |
| p.(Gly256Arg) | 1 | Yes | No | Mild to moderate |
| p.(Gly256Arg) | 2 | NA | No | Mild to moderate |
| p.(Gly256Arg) | 3 | NA | No | Mild to moderate |
| p.(Arg553Trp) | 1 | Yes | Yes | Normal |
| p.(Arg553Trp) | 2 | Yes | No | Normal |
| p.(Arg553Trp) | 3 | No | No | Severe to profound |

Early SCB use corresponds to exposure to treatment during the first month of life.
Early seizure offset corresponds to seizure offset occurring during the first month of life.

SCB: sodium channel blocker
